# Adjusting for Allometric Scaling in ABIDE I Challenges Subcortical Volume Differences in Autism Spectrum Disorder

**DOI:** 10.1101/2020.06.04.20121335

**Authors:** Camille Michèle Williams, Hugo Peyre, Roberto Toro, Anita Beggiato, Franck Ramus

## Abstract

To properly adjust for total brain volume (TBV), brain allometry – the non-linear scaling relationship between regional volumes and TBV – was considered when examining subcortical volumetric differences between typically developing (TD) and Autistim Spectrum Disorder (ASD) individuals.

Autism Brain Imaging Data Exchange I (N = 654) data was analyzed with two methodological approaches: univariate Linear Mixed Effects Models and multivariate Multiple Group Confirmatory Factor Analyses. Analyses were conducted on the entire sample and in subsamples based on age, sex, and Full Scale Intelligence Quotient (FSIQ). A similar ABIDE I study was replicated and the impact of different TBV adjustments on neuroanatomical group differences was investigated.

No robust subcortical allometric or volumetric group differences were observed in the entire sample across methods. Exploratory analyses suggested that allometric scaling and volume group differences may exist in certain subgroups defined by age, sex, and/or FSIQ. The type of TBV adjustment influenced some reported volumetric and scaling group differences.

This study supports the absence of robust volumetric differences between ASD and TD individuals in the investigated volumes when adjusting for brain allometry, expands the literature by finding no group difference in allometric scaling, and further suggests that differing TBV adjustments contribute to the variability of reported neuroanatomical differences in ASD.

---

Autism Spectrum Disorder (ASD) is a neurodevelopmental disorder characterized by early persistent deficits in social communication and interactions and restricted, repetitive patterns of behavior, interests, or activities. These symptoms impair social or occupational functioning and are not restricted to a developmental delay or intellectual deficiencies (American Psychiatric Association, 2013). Although prevalence estimates appear to vary by country and methods of assessments (Adak & Halder, 2017; Elsabbagh et al., 2012; Kim et al., 2011, 2014), ASD prevalence corresponds to 1 child in 59 in the US (Christensen, 2018), an estimate which is consistent with the number of diagnoses reported by parents on national surveys (Kogan et al., 2018). ASD is additionally 3 to 4 times more prevalent in boys than girls (Fombonne, 2009) and is accompanied by an intellectual disability (intelligence quotient, IQ < 70) in one third of patients, while 25% are in the borderline IQ range (from 70 to 85; Christensen et al., 2016). Although diverse genetic (Ramaswami & Geschwind, 2018) and environmental factors (Karimi et al., 2017; Modabbernia et al., 2017), as well as their interactions(Abbott et al., 2018; Rijlaarsdam et al., 2017), are thought to contribute to the complex etiology of ASD, ASD’s etiology remains poorly understood due to the indirect and small effects of known genetic and environmental factors (Crespi, 2016; Varcin et al., 2017). Considering that neuroanatomical markers within the brain are more closely associated to symptoms of a condition, the present study investigated neuroanatomical differences in the Autism Brain Imaging Data Exchange I (ABIDE I, Di Martino et al., 2014; N = 1112) between ASD and typically developing (TD) individuals in terms of their regional (i.e. subcortical and cortical) volumes and the scaling relationship between their regional volumes and Total Brain Volume (TBV; sum of total grey matter (GM) and white matter (WM)).

While toddlers with ASD typically show early brain overgrowth as well as an increase in head circumference (Courchesne, 2002; Hazlett et al., 2005), discrepancies in TBV between ASD and TD individuals after early childhood appear to be relatively subtle - 1–2% greater for ASD (Haar et al., 2016; Riddle et al., 2017)- and to depend on age, intelligence IQ, and sex (Redcay & Courchesne, 2005; Sacco et al., 2015; Stanfield et al., 2008; Sussman et al., 2015). However, since only 20% of autistic individuals experience early brain overgrowth (Zwaigenbaum et al., 2014), global neuroanatomical variation in ASD may reflect a bias in the population norm rather than a trait of ASD (Raznahan et al., 2013). Researchers thus emphasize that TBV differences be examined in light of a population’s inter-individual diversity (Lefebvre et al., 2015; Raznahan et al., 2014) and propose that regional volumes rather than TBV may be better proximal factor candidates underlying ASD (Ecker, 2017).

Numerous Magnetic Resonance Imaging (MRI) studies report neuroanatomical differences between individuals with and without ASD in distributed subcortical and cortical regions thought to contribute to the development of ASD (Ha et al., 2015; D. Y.-J. Yang et al., 2016). For instance, the reduction in GM volume in the hippocampi of children with ASD may feed their episodic memory and social communication impairments (Duerden et al., 2012; Gokcen et al., 2009), and the decrease in GM volume in the superior temporal sulcus and middle temporal gyrus may reflect ASD patients’ social-cognitive deficits (Greimel et al., 2013; Hyde et al., 2010; Wallace et al., 2010). However, reported neuroanatomical group differences in this literature are largely inconsistent and difficult to replicate (Lai et al., 2013; Lenroot & Yeung, 2013; Riddle et al., 2017; Zhang et al., 2018).

The inconsistencies in regional volumetric differences between ASD and healthy individuals are thought to stem from small sample size and heterogeneity, specifically in age (Lin et al., 2015; Riddle et al., 2017; Zhang et al., 2018), sex (Lai et al., 2015, 2017; Mottron et al., 2015; Schaer et al., 2015; Zhang et al., 2018), and IQ variations (Stanfield et al., 2008; Zhang et al., 2018). To address these limitations, researchers are conducting meta-analyses and using cohorts such as the Autism Brain Imaging Data Exchange I (ABIDE I) to investigate the influence of sex, age, IQ, and TBV on brain volumes in ASD. Yet, the conclusions of these studies tend to vary. For example, a meta-analysis examining total and regional brain volume variations across ages in ASD found that the size of the amygdala decreased with age compared to controls (Stanfield et al., 2008), while a recent ABIDE I study reported no such effect and instead identified a smaller putamen in ASD females from 17 to 27 years old (Zhang et al., 2018). Although differences in segmentation algorithms (Katuwal et al., 2016), correction for multiple comparisons, and age range selection may contribute to these discrepancies, studies examining regional neuroanatomical differences in sex (Fish et al., 2017; Jäncke et al., 2015; Mankiw et al., 2017; Reardon et al., 2016, 2018; Sanchis-Segura et al., 2019) and ASD (Lefebvre et al., 2015) report that different methods of adjustment for individual differences in TBV yield varying regional volumetric group differences.

Classical methods of adjustment for TBV (e.g. proportion method (regional volume/TBV), covariate approach) can lead to over and/or under-estimating volumetric group differences (Reardon et al., 2016; Sanchis-Segura et al., 2019) for two reasons. First, they omit the potential group variation in the relationship between a regional volume and TBV. Second, they assume that the relationship between TBV and each regional volume is linear when the relationship is in fact allometric - or non-linear. If the relationship between TBV and a regional volume was linear, the exponent (α) of the power equation: 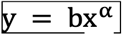 would be equal to 1, indicating isometry. However, the exponent tends to be either hyperallometric (α > 1) or hypoallometric (α < 1) depending on the regional volume (Finlay et al., 2001; Mankiw et al., 2017; Reardon et al., 2016, 2018). When a region shows an hypoallometric coefficient, the regional volume increases less than TBV as TBV increases and when the coefficient is hyperallometric, the regional volume increases more than TBV as TBV increases (e.g. Liu, Johnson, Long, Magnotta, & Paulsen, 2014; Mankiw et al., 2017).

Adjusting for differences in TBV with allometric scaling has two major implications for neuroanatomical research in ASD. First, if the allometric coefficient (α) differs between individuals with and without ASD, the relationship between regional and total volumes may serve as an additional cerebral marker to differentiate between groups. Second, allometric scaling group differences aside, adjusting for the allometric relationship of each subcortical and cortical volume with total volume yields a more precise estimate of each regional volume, and in turn, provides a more accurate evaluation of volumetric group differences.

To this day, brain allometry in ASD has only been considered in two studies that examined corpus callosum and cerebellar differences between ASD and control individuals (Lefebvre et al., 2015; Traut et al., 2018, respectively). Thus, the primary goal of this study was to investigate allometric scaling and volumetric differences between ASD and control individuals in subcortical volumes while taking into account brain allometry. The second aim was to identify whether neuroanatomical group differences depend on sex, age and/or FSIQ, variables previously reported to influence group differences in brain volumes in studies where brain allometry was omitted (Stanfield et al., 2008; Zhang et al., 2018). As the first study to investigate and adjust for allometric scaling differences in local volumes between TD and ASD individuals, no a priori hypotheses were postulated.

Subcortical allometric and volumetric group differences were investigated in the ABIDE I, a cohort which consists of 539 individuals with ASD and 573 age and sex matched controls (A. Di Martino et al., 2014). A Multiple Group Confirmatory Factor Analysis (MGCFA), a multivariate statistical approach which advantageously tests for global group differences in brain allometry and considers the mutual relationship between regional brain structures (de Jong et al., 2017; Toro et al., 2009) was conducted on the entire sample and subsamples based on age, sex, and FSIQ. However, considering the recency of the MGCFA to examine volumetric group differences (de Mooij et al., 2018; Peyre et al., 2020), results from the MGCFA were compared to those obtained from Linear Mixed Effects Models (LMEMs). The present study additionally attempted to replicate the age and sex subcortical differences Zhang and colleagues’ (2018) found in the ABIDE I without adjusting for brain allometry and examined how four different TBV adjustment techniques influence the replicated results.

## Methods

### 1. Participants

#### Participant Recruitment

Data was obtained from ABIDE I: a consortium with 1112 existing resting-state functional magnetic resonance imaging datasets with corresponding structural MRI and phenotypic information on 539 ASD patients and 573 age-matched controls between 6 to 64 years old from 17 different scanner sites (http://fcon_1000.projects.nitrc.org/indi/abide; Di Martino et al., 2014). ASD individuals were diagnosed by (1) clinical judgment only, or (2) using the Autism Diagnostic Observation Schedule (ADOS) and/or Autism Diagnostic Interview–Revised, or by (3) combining clinical judgment and the diagnostic instruments only. Di Martino and collegues (2014) reported that 94% of the 17 sites using the ADOS and/or Autism Diagnostic Interview-Revised obtained research-reliable administrations and scorings. Data was anonymized and collected by studies approved by the regional Institutional Review Boards. Further details on participant recruitment and phenotypic and imaging data analyses are provided by Di Martino and colleagues (2014).

#### Exclusion/Inclusion Criteria

As in Zhang and colleagues’ (2018) study that we aimed to replicate, individuals over 27 years old when scanned were excluded from the analyses since the age distribution was skewed to the left and subjects over 27 years old had a broad age distribution. Moreover, participants with an FSIQ or linearly estimated FSIQ by Lefebvre and colleagues (2015; details in their Supp. Info. *<u>Intelligence Score</u>*) greater than 70 and smaller than 130 were included in the analyses to create a more homogenous sample.

Finally, participants were included based on the visual quality checks that were performed on the Freesurfer v.5.1 segmentations *(http://surfer.nmr.mgh.harvard.edu/)*. Indeed, considering that segmentation errors can yield large volume estimation errors, we decided to use the stringent image and segmentation quality criteria that was also applied by Lefebvre and colleagues (2015) at the cost of a reduction of sample size. Considering that the same segmentation and quality check standard was not available for ABIDE II (Adriana Di Martino et al., 2017) or for cortical regions, cortical ABIDE I data and ABIDE II data were not included in this study.

Given that 3 controls and 33 ASD individuals exhibited differing comorbidities (e.g. Attention Hyperactivity Deficit Disorder, Obsessive Compulsive Disorder, phobias (e.g. spiders, darkness)) varying in severity, all individuals with comorbidities were maintained in the main analyses and were removed from the post-hoc analyses performed without outlier values to consider their impact on reported group differences.

#### Entire Sample’s Descriptive Statistics

The entire sample consisted of 654 participants (302 ASD and 352 controls) following the Freesurfer v.5.1 segmentation quality checks. The 302 ASD and 352 TD individuals differed in terms of sex ratio and FSIQ but not in handedness or age (Table 1). There were 218 ASD participants with a total ADOS score (M = 11.85, SD = 3.76), which corresponds to the sum of the ADOS communication subscore (M = 3.72, SD = 1.49) and the ADOS social interactions subscore (M = 8.15, SD = 2.72).

**Table 1.** Descriptive Statistics of the Entire Sample in Sex, Age, Handedness, ADOS, and FSIQ.

|  | ASD<br>(N = 302) | TD<br>(N = 352) | Statistics |  |
| --- | --- | --- | --- | --- |
| Sex Ratio (M/F) | 265/37 | 283/69 | $\chi^2 (1) = 6.47$ | p = 0.012 |
| Age in Years (SD) |  |  |  |  |
| Mean | 14.54 (4.47) | 14.54 (4.55) | $\chi^2 (1) = 0.02$ | p = 0.877 |
| Min | 7.00 | 6.47 |  |  |
| Max | 26.95 | 26.85 |  |  |
| Handedness (Right/Other) | 170/30 | 223/24 | $\chi^2 (1) = 2.43$ | p = 0.119 |
| FSIQ |  |  |  |  |
| Mean (SD) | 102.18 (14.37) | 109.75 (11.05) | $\chi^2 (1) = 48.58$ | p < 0.001 |
| Min | 71.00 | 73.00 |  |  |
| Max | 129.11 | 129.00 |  |  |
| ADOS Total |  |  |  |  |
| Mean (SD) | 11.85 (3.76) |  |  |  |
| Min | 2 |  |  |  |
| Max | 21 |  |  |  |
| ADOS Communication |  |  |  |  |
| Mean (SD) | 3.72 (1.49) |  |  |  |
| Min | 0 |  |  |  |
| Max | 7 |  |  |  |
| ADOS Social Interactions |  |  |  |  |
| Mean (SD) | 8.15 (2.72) |  |  |  |
| Min | 2 |  |  |  |
| Max | 14 |  |  |  |

*N.B.* Standard deviation in parentheses. Other: Left, Ambidextrous, or Mixed. FSIQ: Full Scale Intelligence Quotient. ASD: Autism Spectrum Disorder. TD: Typically Developing. M: Male. F: Female. Handedness was only provided for a subset of individuals. $\chi^2$ from the Kruskal-Wallis test for FSIQ and Age. ADOS (Autism Diagnostic Observation Schedule) Total corresponds to the sum of the ADOS communication and ADOS social interactions scores.

#### Subsamples’ Descriptive Statistics

In addition to the analyses on the entire sample, we ran exploratory MGCFAs and LMEMs on four sufficiently powered subsamples (Mundfrom et al., 2005) to investigate age, sex, and FSIQ interactions which cannot be simultaneously investigated with the MGCFA. Girls (N_ASD_ = 37, N_Control_ = 69) and adults aged 20 to 27 years old (N_ASD_ = 46, N_Control_ = 54) could not be examined in the subsample analyses due to the insufficient number of participants (N < 50; Mundfrom et al., 2005).

Subgroups were additionally defined based on previous studies reporting age effects in ASD (e.g. Lin et al., 2015; Stanfield et al., 2008; Zhang et al., 2018): boys aged 6 to under 12 years old (N_ASD_ = 87, N_Control_ = 97) and boys aged 12 to under 20 years old (N_ASD_ = 138, N_Control_ = 141). Age did not differ between ASD and TD individuals in each group.

In light of the group differences in FSIQ and the association between FSIQ and brain volume (Maier et al., 2015; McDaniel, 2005), subsamples were additionally created based on the boys’ median FSIQ, yielding boys with an FSIQ ≤ 107.8 (N_ASD_ = 165, N_Control_ = 109) and boys with an FSIQ > 107.8 (N_ASD_ = 100, N_Control_ = 174). FSIQ was smaller in ASD (M = 93.40, SE = 0.77) compared to control boys with an FSIQ ≤ 107.8 (M = 98.96, SE = 0.72; ß = -0.58, SE = 0.12, p = 1.4 × 10^-06^). FSIQ did not differ across boys with an FSIQ > 107.8 (ß = -0.01, SE = 0.13, p = 0.952).

Further descriptive statistics on brain volumes, age, FSIQ score by group and sex were reported for the entire sample and descriptive statistics on brain volumes, age, FSIQ score by group were reported for each subsample (Tables *S1*-*S4*) as well as the distribution of all brain volumes, age, and FSIQ of Autism Spectrum Disorder (ASD) and control participants in Figures S1-13 to compare to those from Zhang and colleagues’ (2018) study. Finally, since the sample size was predefined, power analyses were run *a posteriori* on significant LMEM main effects and interactions with the simr package (Green & MacLeod, 2016; Supp Info 2. *Power Analyses*).

### 2. Analyses

Analyses performed on R (R Core Team, 2019) were preregistered ahead of time on OSF (https://osf.io/wun7s), except where indicated. The data and scripts that support the findings and figures of this study are openly available in “Subcortical-Allometry-in-ASD” at http://doi.org/10.5281/zenodo.3592884.

Since previous research either did not examine the left and right hemisphere scaling coefficients of some of the presently investigated volumes (Jong et al., 2017; Liu et al., 2014; Reardon et al., 2016), we examined whether there were allometric (non-linear) relationships between regional and total brain volumes, in ASD and TD individuals separately. Although not preregistered, we reported scaling coefficients with the 95% confidence interval and tested whether the scaling coefficients of each regional region with TBV differed from 1 with the car R package (Fox & Weisberg, 2019). Analyses were conducted with and without age, sex, and age by sex interactions to examine the extent to which these additional variables influence the scaling coefficients. Additional analyses were also conducted without outliers, without individuals with comorbidities, and with medication use (medication *versus* no medication) as a covariate to assess whether scaling coefficients were robust to these factors.

MGCFAs and LMEMs were conducted to address the study’s primary goal to investigate allometric scaling and volumetric group differences and secondary goal to examine whether allometric scaling and volumetric group differences depend on age, sex and or FSIQ. Briefly, a MGCFA is a multivariate approach that involves simultaneous confirmatory factor analyses (CFA) in two or more groups and tests measurement invariance across groups (i.e. that the same model of equations measures the same latent construct). In a CFA, observed variables (brain volumes) are used to measure an unobserved or latent construct (TBV). A CFA in turn corresponds to a system of equations that describes the relationship the observed variables and the latent construct they measure (TBV). MGCFAs advantageously measure group (i.e. ASD versus Control) differences across all regional volumes simultaneously (i.e. global test) and in each regional volume (i.e. regional test), while adjusting for mutual relationships between regional brain volumes. MGCFAs were run with the lavaan R package (Rosseel, 2012). We additionally conducted LMEMs, which measure group differences in each regional volume separately, with the lmerTest R package (Kuznetsova et al., 2017) to (i) evaluate the consistency between MGCFA and LMEMs results; (ii) adjust covariates that could not be included in MGCFAs; and (iii) facilitate result comparisons with previous studies examining neuroanatomical differences in ASD that conducted LMEMs.

#### Equations in the MGCFA

The observed variables estimating the latent construct (i.e. TBV) were the following 22 regional volumes (Table 2). All brain volumes were log10 transformed in order to take into account the power relationship between each regional volume and TBV within the general linear model framework. This yielded the linear allometric scaling equation (1) where i corresponds to the investigated regional volume, α to the exponent of the power relationship (the allometric coefficient), and group to ASD or Control:

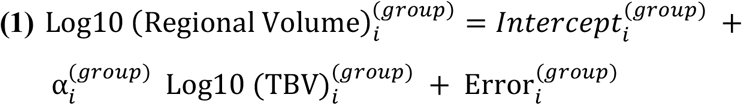

**Table 2.** Investigated Regional Volumes.

| Total Cerebral White Matter |  |
| --- | --- |
| Brain-Stem |  |
| Right Ventral Diencephalon | Left Ventral Diencephalon |
| Right Cerebellum Cortex | Left Cerebellum Cortex |
| Right Accumbens | Left Accumbens |
| Right Amygdala | Left Amygdala |
| Right Caudate | Left Caudate |
| Right Hippocampus | Left Hippocampus |
| Right Pallidum | Left Pallidum |
| Right Putamen | Left Putamen |
| Right Thalamus Proper | Left Thalamus Proper |
| Right Hemisphere Cortex | Left Hemisphere Cortex |

#### Testing Measurement Invariance in the MGCFA

First, TBV differences between groups identified by regressing TBV on group in the MGCFA models were adjusted for in the configural models of each sample. Second, configural invariance – whether the same observed variables explain the same latent construct across groups - was tested by establishing a configural model with correlated residuals between regional volumes that similarly fits both groups when the intercept and slope values of the allometric equations for each regional volume differs between ASD and Controls. Good model fit was determined using commonly used fit indices: the Tucker Lewis Index (TLI), the Comparative Fit Index (CFI), and the Root Mean Square Error of Approximation (RMSEA) with a TLI and CFI > .95 and a RMSEA ≤ .06 indicating good fit (Hu & Bentler, 1999). TLI, CFI, and RMSEA robust fit indices were used to correct for non-normality and were obtained from the Maximum Likelihood Robust (MLR) estimator from the lavaan package (Rosseel, 2012). Although we preregistered that we would additionally use the Standardized Root Mean Square Residual (SRMR), the SRMR was not used since the lavaan package (Rosseel, 2012) does not provide a robust SRMR.

Third, allometric scaling group differences were identified by testing for metric invariance (equality of slopes, or α_i_ coefficients from Equation 1) between groups and fourth, volumetric group differences adjusted for allometric scaling were identified by testing for scalar invariance (equality of intercepts, or Intercepts from Equation 1) between groups. Metric and scalar invariance are tested with a global test followed by a regional test in each volume if the global test is significant. In a global metric invariance test, regional volumes are simultaneously tested for allometric scaling (slope) group differences by comparing the configural model where the intercept and slope values differ between groups to a model where the slope values are constrained (the same) across groups. In a global scalar invariance test, regional volumes are simultaneously tested for volumetric (intercept) group differences by comparing the configural model where the intercept values differ between groups to a model where intercept values (and slope values, if global metric invariance is rejected) are constrained across groups. If the metric and/or scalar global invariance test is significant (χ^2^ difference test p-value < 0.05) and robust TLI, CFI, and RMSEA indicate better model fit for configural model (F. Chen et al., 2008; Hu & Bentler, 1999), groups respectively differ in allometric scaling (slopes) and/or volumes (intercept) in one or more of the regional volumes.

Regional volumes that differ in terms of allometric scaling and/or volume between groups are then identified by conducting a regional invariance test on each volume. In a regional invariance test, a model where the parameter (e.g. intercept, slope) values are constrained across groups is compared to a model where all but one of the parameter values of a regional volume are constrained across groups. We initially preregistered the following criteria for significant group differences in parameters in regional invariance tests based on the literature on confirmatory factor analyses. Groups would differ in parameter value if the χ^2^ test differed between groups was significant, if the p value was below 0.05 / 22 (Bonferroni correction for the number of regional volumes in the MGCFA), and if the group difference in model fit exceeded Chen’s (2008) cutoffs (|∆CFI| >.005 & robust |∆RMSEA| ≥.010 for unequal samples with under 300 participants). These more conservative cutoffs were chosen since our entire sample ASD group (N = 302) neared the 300 participants mark. However, these criteria lead to a mismatch between the MGCFA and LMEMs.

As a result, since there is currently no rule of thumb for the cutoff values of fit indices that should be employed in varying conditions (e.g. number of observed variables and factors), leaving researchers to choose fit criteria (Putnick & Bornstein, 2016), we considered there to be a neuroanatomical group difference, even if Chen’s (2008) criteria indicated invariance, if (i) the χ^2^ difference test was significant, (ii) the MGCFA effect size was greater than 0.2, (iii) the corresponding LMEM (see next paragraph) effect size was similar to the MGCFA effect size, and (iv) the corresponding LMEM with False Discovery Rate correction (FDR) was significant with and without outliers.

#### LMEMs

Corresponding LMEMs were run on regional volumes in the entire sample and on volumes that significantly differed between ASD and control participants in terms of allometry (as indicated by the regional metric invariance test) and/or volume (as indicated by the regional scalar invariance test) in the exploratory subsamples. TBV was calculated as the sum of grey and white matter. The same equation used in the MGCFA was entered in the LMEMs except that scanner site was also included in the equation (2) as random intercept (slopes were not written out in equation (2) for clarity).

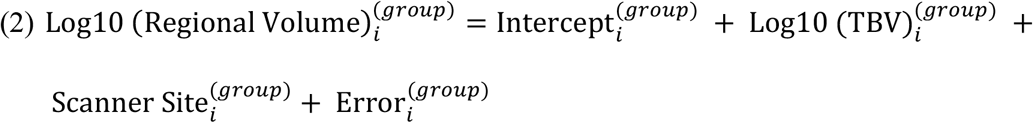

An interaction of log10(TBV) by group indicated a significant difference in allometric scaling between groups while a significant group effect suggested a significant volumetric group difference. Additional sensitivity analyses were run excluding outliers and individuals with comorbidities, and with medication use as a covariate to ensure that the findings were robust.

#### Testing Age, Sex, & FSIQ Effects

To address the study’s secondary goal, which is to identify whether allometric scaling and/or volumetric group differences depend on the effects of age, sex, and/or FSIQ, we ran LMEMs in the entire sample with equation (3) where scanner site was included as a random intercept (slopes were not written out in equation (3) for clarity).

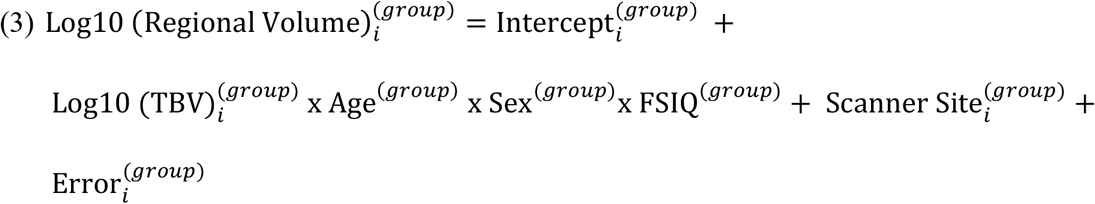

#### Testing Age, Sex, & FSIQ Effects: Exploratory Analyses

We ran several exploratory MGCFAs and LMEMs on four boy subsamples to further compare MGCFA and LMEM results. In each boy subsample, we applied equation (1) in the exploratory MGCFAs and equation (2) in the exploratory LMEMs for each regional volume. Scanner site was always included as a random effect in the LMEMs. If groups in a subsample differed in terms of age and/or FSIQ, we ran LMEMs in the subsamples with age and/or FSIQ as interactive fixed effects (i.e. if groups differed in terms of age, the group × log10(TBV) × age interaction was included) on the volumes that were and were not identified by the MGCFA and corresponding LMEMs as exhibiting group differences. We conducted LMEMs on volumes that were not identified as exhibiting group differences according to the MGCFA and corresponding LMEMs to investigate if some neuroanatomical group differences are only apparent when considering age and/or FSIQ effects. Total GM volume was also investigated in the latter LMEMs. All possible interactions were maintained in all LMEMs and only significant main effects and interactions were reported. LMEMs revealing significant neuroanatomical group differences were also conducted without outliers, individuals with comorbidities, and with medication use as a covariate to ensure that the findings were robust.

#### Testing the Relationship of ASD severity with Neuroanatomical Group Differences

We additionally ran post-hoc analyses, which were not preregistered, on brain regions exhibiting neuroanatomical group differences to examine whether the total ADOS score in ASD individuals was a significant predictor of the investigated volume and the allometric scaling relationship between that volume and TBV. These LMEMs were run on ASD individuals with the same fixed and random effects as the LMEMs revealing neuroanatomical group differences, except that the Group fixed effect was replaced by the total ADOS score. LMEMs were additionally conducted without outliers and individuals with comorbidities and medication status was added as a covariate. Other ASD scores available in ABIDE I were not employed due to the small number of individuals in each category (Supp Info 3. *MGCFA & LMEMs Assumptions*).

#### Testing the Influence of TBV Adjustment Techniques on Reported Neuroanatomical Differences

The additional LMEMs, which were run to address the study’s third goal to contribute to the literature suggesting that neuroanatomical group differences vary depending on the applied TBV adjustment technique, were not preregistered. We examined the influence of 4 types of TBV adjustment techniques by comparing results from LMEMs (i) without TBV adjustment (e.g. Zhang et al., 2018), (ii) with a linear adjustment considering TBV as a covariate (most common; Prigge et al., 2013; van Rooij et al., 2017; Zhang et al., 2018), (iii) with linear adjustment while considering the interaction of TBV by Group (e.g. Lefebvre et al., 2015), and (iv) with an allometric scaling adjustment by considering the interaction of log10(TBV) by Group (e.g. Lefebvre et al., 2015; Mankiw et al., 2017; Sanchis-Segura et al., 2019). In the no adjustment and linear adjustment LMEMs, all volumes were standardized raw volumes.

#### Testing the Influence of TBV Adjustment Techniques on our Replication of Zhang and Colleagues’ (2018) Study

In a second attempt to address the study’s third goal, we sought to replicate the study by Zhang and colleagues (2018), who similarly examined the subcortical correlates of ASD with ABIDE I, as to examine the influence of different adjustment techniques on the findings that we successfully replicated.

Dependent variables in the LMEMs were the Cortical WM Volume, Total GM Volume, the caudate, the amygdala, the hippocampus, the thalamus, the pallidum, the putamen, and the accumbens. Scanner site was always included as a random intercept and subject as a random intercept when hemisphere was included in the LMEMs. Fixed effects differed based on the type of adjustment technique, as described below. Dependent and independent variables were entered in the models as raw values except for age (linear and quadratic), which was centered (i.e. demeaned). Significant group main effects and interactions were reported and compared across LMEMs with varying adjustment techniques and p values were not adjusted for multiple comparison as in Zhang and colleagues’ (2018) study.

#### LMEMs without TBV Adjustment

Fixed effects were sex, age (quadratic or linear), hemisphere (except for Cerebral WM and Total GM volumes), and group (ASD and Controls). Two replication strategies were put into place: a “result replication” and a “methodological replication”. In the “result replication”, models were identified based on the significant interactions reported by Zhang and colleagues (2018) to compare effect sizes even if group interactions and main effects were not statistically significant in our sample. In the “methodological replication”, LMEMs were identified using Zhang and colleagues’ (2018) technique of maintaining main effects in the model and sequentially removing non-significant interactions (p > 0.05) from the model.

#### LMEMs with linear TBV Adjustment

As in Zhang and colleagues (2018) analyses, TBV was added as a covariate to the LMEMs identified with the “result replication” and “methodological replication” techniques. Although the authors commented on whether results were similar after covarying for TBV, they did not provide statistics (i.e. effect sizes, p values).

#### Comparing LMEMs with the lack of and differing TBV Adjustment Techniques

All brain volumes were log10 transformed prior to scaling. LMEMs identified with the “result replication” and “methodological replication” techniques were run with the interaction of group by log10 (TBV).

## Results

### I- Testing for Allometry

When examining the relationship of each regional volume with TBV, we found that cerebral white matter was hyperallometric (slope >1), the cortical volume was isometric (slope = 1), and most subcortical regions were hypoallometric (slope < 1). After removing outliers and including medication as a fixed effect, all subcortical regions were hypoallometric except for the right amygdala in controls which remained isometric (*α* = 0.74, CI low = 0.73, CI high = 1.02, p = 0.094; Tables S6 - S12).

When examining the relationship of each regional volume with TBV and considering the interaction of age by sex, we found that cerebral white matter was hyperallometric (slope >1), cortical volume was isometric (slope = 1), and most subcortical regions were hypoallometric (slope < 1). After removing outliers and including medication as a fixed effect, all subcortical regions were hypoallometric except for the right amygdala in controls which remained isometric (*α* = 0.86, CI low = 0.71, CI high = 1.02, p = 0.094; Tables S10 - S13). Medication use was not significant across regional volumes for ASD and Control individuals.

### II- Global Allometric and Volumetric Group Differences

In the MGCFA, the variance of TBV (the latent factor) was set to one to freely estimate the factor loading of the first regional volume. As a result, all ß reported from the MGCFA correspond to standardized effect sizes where the variance of regional volume and TBV are set to 1. Group differences in the MGCFA were estimated by calculating the group difference in standardized slopes and intercepts.

In the LMEMs, standardized estimates, ß, were reported by centering and scaling dependent and independent variables. Reported p-values are not corrected for multiple comparisons in the MGCFAs and were FDR corrected for the LMEMs. Statistics were reported for the age measure (age or age^2^) with the largest effect size estimate. Correlated residuals slightly differed across samples (Table S14) and MGCFA model fit were acceptable (Table S15; Supp Info *4.MGCFA Results*).

Since factor levels were set to 1: Controls and 2: ASD in all LMEMs conducted in this study, a negative effect size in the MGCFA suggests that the slope or intercept is greater for Controls compared to ASD individuals, while a positive effect size suggests that the slope or intercept is smaller for Controls compared to ASD individuals.

#### TBV Group Differences

TBV did not differ between individuals with and without ASD in the entire sample (ß = 0.03, SE = 0.06, p = 0.431) in the MGCFA or LMEM (ß = -0.01, SE = 0.07, p = 0.878).

#### Global Allometric Scaling Group Differences

Global metric invariance was supported in the entire sample (∆χ2_(22)_ = 17.4, p = 0.7395), suggesting that there was no allometric scaling (slope) difference between ASD and TD individuals.

#### Global Volumetric Group Differences

Scalar invariance was supported in the entire sample (∆χ2_(22)_ = 26.1, p = 0.2487), suggesting that there are no regional volumetric differences between ASD and TD individuals when adjusting for individual differences in TBV by taking into account allometric scaling.

#### LMEMs

LMEMs were consistent with the MGCFA except for a group effect found in the right pallidum (ß = 0.15, SE = 0.06, p = 0.028). This group effect was no longer significant (ß = 0.07, SE = 0.06, p = 0.426) after removing outliers and individuals with comorbidities and controlling for medication use.

### III- Dependence of Allometric Scaling and/or Volumetric Group Differences on Age, Sex, and/or FSIQ Effects

Only significant results are reported (see Supp Info 4. *MGCFA Results*).

#### TBV Group Differences

MGCFAs only revealed a group difference in TBV for boys with an FSIQ ≤ median (107.8) where ASD individuals had a greater TBV than controls (ß = 0.13, SE = 0.09, p = 0.023). Results from the LMEMs were consistent with those of the MGCFA. There was a significant interaction of sex by group by FSIQ in the entire sample (ß = -0.52, SE = 0.21, p = 0.048), which was due to the greater TBV in ASD boys with an FSIQ ≤ median (M = 1 218.37 cm^3^, SE = 0.76 cm^3^) compared to their control counterparts (M = 1 181.89 cm^3^, SE = 1.09 cm^3^; ß = 0.23, SE = 0.11, p = 0.027). Non-significant TBV group differences are provided as Supp Info 4. *MGCFA Results*.

#### Global Allometric Scaling Group Differences

Global metric invariance was supported in boys from 6 to under 12 years old (∆χ2_(22)_ = 22.9, p = 0.405) and in boys with an FSIQ ≤ 107.8 (∆χ2_(22)_ = 20.0, p = 0.586), suggesting that there was no allometric scaling (slope) difference between ASD and TD individuals in these samples. However, global metric invariance was not supported in boys from 12 to under 20 years old (∆χ2_(22)_ = 38.7, p = 0.015) and in boys with an FSIQ > 107.8 (∆χ2_(22)_ = 38.5, p = 0.016). Thus, a regional metric invariance test was conducted on each regional volume of these subsamples to establish where allometric scaling discrepancies between groups lied.

#### Regional Allometric Scaling Group Differences in Boys Aged 12 to under 20 years old

Regional metric invariance χ^2^ difference test indicated that the constrained configural model significantly differed from the constrained configural model with one freed slope, when the slope was freed for the brain stem (ß = -0.06, ∆χ2_(1)_ = 11.7, p = 6.13 × 10^-3^), the left amygdala (ß = 0.08, ∆χ2_(1)_ = 11.3, p = 7.87 × 10^-4^), and the right hippocampus (ß = 0.22, ∆χ^2^_(1)_ = 58.2, p = 2.34 × 10^-14^). Although the robust CFI and robust RMSEA fit indices were invariant across models according to Chen’s metric invariance cutoffs (|∆CFI| >.005 & |∆RMSEA| ≥.010; Table S16.A), the present study’s four step procedure for determining invariance suggested that the allometric scaling relationship between the right hippocampus and TBV differed between groups. ASD boys aged 12 to under 20 years old had a smaller allometric scaling coefficient (ß = 0.52, SE = 0.01, p = 2.21 × 10^-8^) than their control counterparts (ß = 0.74, SE = 0.01, p = 2.40 × 10^-8^).

Specifically, the χ^2^ difference test indicated a group difference in the right hippocampus, the group difference (ß = 0.22) was greater than 0.2, the allometric scaling group difference was replicated in the corresponding LMEM with (Figure S17) and without (Figure 1) outliers and the effect size of the MGCFA and LMEMs were similar (Table 3.A &B).

**Figure 1.**
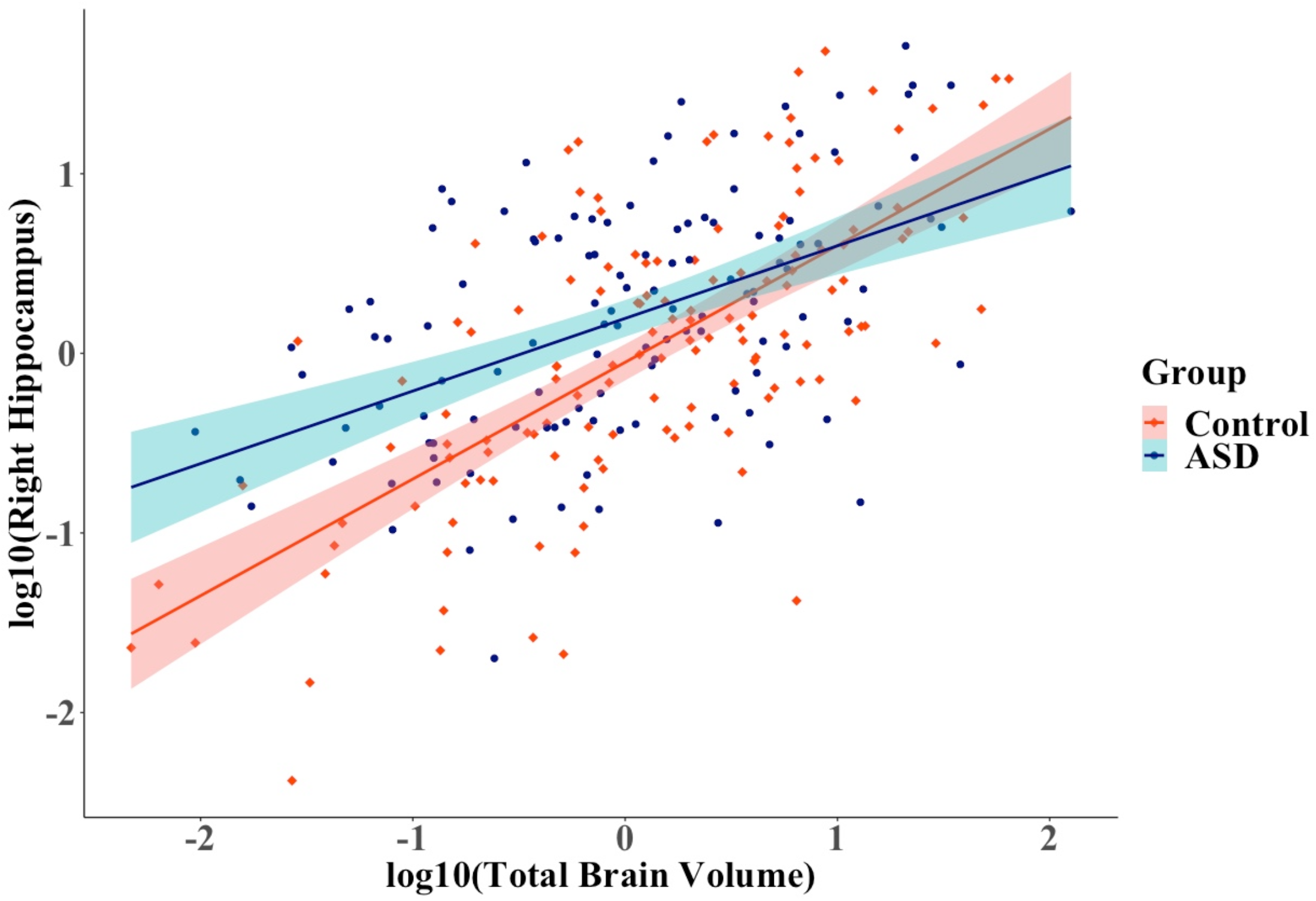
Relationship between the right hippocampus and total brain volume across groups after outlier and comorbidity removal (N Control = 137, N ASD =123) in boys aged 12 to under 20 years old. ASD, Autism Spectrum Disorder. 95% confidence region are given by group. Volumes were log transformed and scaled.

**Table 3.** Right Hippocampus LMEM Results (A) and Unstandardized Allometric Coefficients (B) for Boys from 12 to 20 years old.

| A) | **Right Hippocampus ~ Group x log10(TBV)** |  |  |  |  |  |
| --- | --- | --- | --- | --- | --- | --- |
|  | (N<sub>ASD</sub> = 138<br>& N<sub>C</sub> = 141) |  |  | No Outliers & Comorbidities<br>(N<sub>ASD</sub> = 123 & N<sub>C</sub> = 137) |  |  |
| | $\beta$ | SE | pFDR | $\beta$ | SE | pFDR |
| Medication |  |  |  | -0.03 | 0.12 | 0.855 |
| log10(TBV) | 0.83 | 0.07 | $1.76 \times 10^{-24}$ | 0.65 | 0.06 | $4.48 \times 10^{-23}$ |
| Group | 0.22 | 0.09 | 0.020 | 0.21 | 0.08 | 0.010 |
| Group x<br>log10(TBV) | -0.33 | 0.10 | 0.003 | -0.25 | 0.08 | 0.010 |

| B) | **Right Hippocampus ~ Group x log10(TBV)** |  |  |  |  |  |
| --- | --- | --- | --- | --- | --- | --- |
|  | (N<sub>ASD</sub> = 138<br>& N<sub>C</sub> = 141) |  |  | No Outliers & Comorbidities<br>(N<sub>ASD</sub> = 123 & N<sub>C</sub> = 137) |  |  |
| | $\alpha$ | SE | pFDR | $\alpha$ | SE | pFDR |
| log10(TBV) |  |  |  |  |  |  |
| ASD | 0.66 | 0.09 | 0.000 | 0.53 | 0.09 | 0.000 |
| Control | 1.08 | 0.10 | 0.000 | 0.82 | 0.08 | 0.000 |

*N.B.* $\beta$ corresponds to standardized beta for all main effects and interactions. $\alpha$ corresponds to the unstandardized allometric scaling coefficient of log10(TBV) with log10(left Accumbens). C corresponds to controls, FDR to false discovery rate correction for multiple comparison, TBV to Total Brain Volume and FSIQ to Full Scale Intelligence Quotient.

To examine if the allometric scaling group difference reported the right hippocampus of boys from 12 to under 20 years old depended on FSIQ, we ran a LMEM with the right hippocampus as dependent variable, TBV by group by FSIQ as fixed effects and scanner site as random intercept. Again, ASD individuals had a smaller allometric scaling coefficient compared to controls before and after outlier and comorbidity removal and medication use inclusion (Table 4. A & B; *a posteriori* Power Analyses Table *S17*).

Post hoc analyses revealed that the total ADOS score did not significantly predict right hippocampal volume (ß = 10.01, SE = 0.01, p = 0.695) or the allometric scaling relationship (ß = 0.01, SE = 0.02, p = 0.695) of that volume in ASD individuals with an available total ADOS score (N = 81).

**Table 4.** Right Hippocampus LMEM Results (A) and Unstandardized Allometric Coefficients (B) for Boys from 12 to 20 years old.

|  |  |  |  |  |  |  |
| --- | --- | --- | --- | --- | --- | --- |
| A) | **Right Hippocampus ~ Group x log10(TBV) x FSIQ** |  |  |  |  |  |
|  | (N<sub>ASD</sub> = 138<br>& N<sub>C</sub> = 141) |  |  | No Outliers & Comorbidities<br>(N<sub>ASD</sub> = 119 & N<sub>C</sub> = 133) |  |  |
| | $\beta$ | SE | pFDR | $\beta$ | SE | pFDR |
| Medication |  |  |  | -0.04 | 0.11 | 0.739 |
| log10(TBV) | 0.86 | 0.08 | 9.34 x10<sup>-22</sup> | 0.69 | 0.06 | 2.71 x 10<sup>-23</sup> |
| Group | 0.22 | 0.10 | 0.059 | 0.21 | 0.09 | 0.021 |
| FSIQ | -0.09 | 0.08 | 0.503 | 0.03 | 0.06 | 0.739 |
| Group x log10(TBV) | -0.35 | 0.11 | 0.004 | -0.22 | 0.09 | 0.037 |
| Group x FSIQ | 0.08 | 0.10 | 0.503 | -0.08 | 0.07 | 0.537 |
| FSIQ x log10(TBV) | 0.06 | 0.07 | 0.503 | 0.03 | 0.06 | 0.739 |
| Group x FSIQ x<br>log10(TBV) | -0.02 | 0.10 | 0.816 | 0.06 | 0.08 | 0.739 |
| B) | **Right Hippocampus ~ Group x log10(TBV) x FSIQ** |  |  |  |  |  |
|  | (N<sub>ASD</sub> = 138<br>& N<sub>C</sub> = 141) |  |  | No Outliers & Comorbidities<br>(N<sub>ASD</sub> = 119 & N<sub>C</sub> = 133) |  |  |
| | $\alpha$ | SE | pFDR | $\alpha$ | SE | pFDR |
| log10(TBV) |  |  |  |  |  |  |
| ASD | 0.67 | 0.09 | 0.000 | 0.62 | 0.09 | 0.000 |
| Control | 1.11 | 0.11 | 0.000 | 0.86 | 0.08 | 0.000 |

*N.B.* $\beta$ corresponds to standardized beta for all main effects and interactions. $\alpha$ corresponds to the unstandardized allometric scaling coefficient of log10(TBV) with log10(left Accumbens). C corresponds to controls, FDR to false discovery rate correction for multiple comparison, TBV to Total Brain Volume and FSIQ to Full Scale Intelligence Quotient.

#### Regional Allometric Scaling Group Differences in Boys with an FSIQ > median (107.8)

The constrained configural model with one freed slope significantly differed from the constrained configural model, when the slope was freed for the left hippocampus (ß = 0.11, ∆χ^2^ _(1)_ = 9.1, p = 0.003), the left caudate (ß = 0.04, ∆χ2_(1)_ = 4.84, p = 0.027), the left accumbens (ß = 0.21, ∆χ2_(1)_ = 6.2, p = 0.013), left pallidum (ß = 0.22, ∆χ2_(1)_ =7.8, p = 0.005), and the right ventral diencephalon (ß = 0.05, ∆χ2_(1)_ = 5.9, p = 0.015). Since the covariance matrix of the residuals was not positive definite in group 2, we were not able to interpret the cortical white matter freed slope model. Although the robust CFI and robust RMSEA fit indices were invariant across models according to Chen’s metric invariance cutoffs (|∆CFI| >.005 & |∆RMSEA| ≥.010; Table S16.B, the present study’s four step procedure for determining invariance supports that the allometric scaling relationship between the left accumbens and TBV differed between groups. ASD boys with an FSIQ > median having a smaller allometric scaling coefficient (ß = 0.32, SE = 0.01, p = 2.13 × 10^- 3^) than their control counterparts (ß = 0.52, SE = 0.02, p = 3.76 × 10^- 3^). Specifically, the χ2 difference test indicated a group difference in the left accumbens, the group difference (× = 0.21) was greater than 0.2, the allometric scaling group difference was replicated in the corresponding LMEM with (Figure S21) and without outliers (Figure 2) and the effect size of the MGCFA and LMEMs were similar (Table 5. A &B; *a posteriori* Power Analyses Table *S17*).

**Figure 2.**
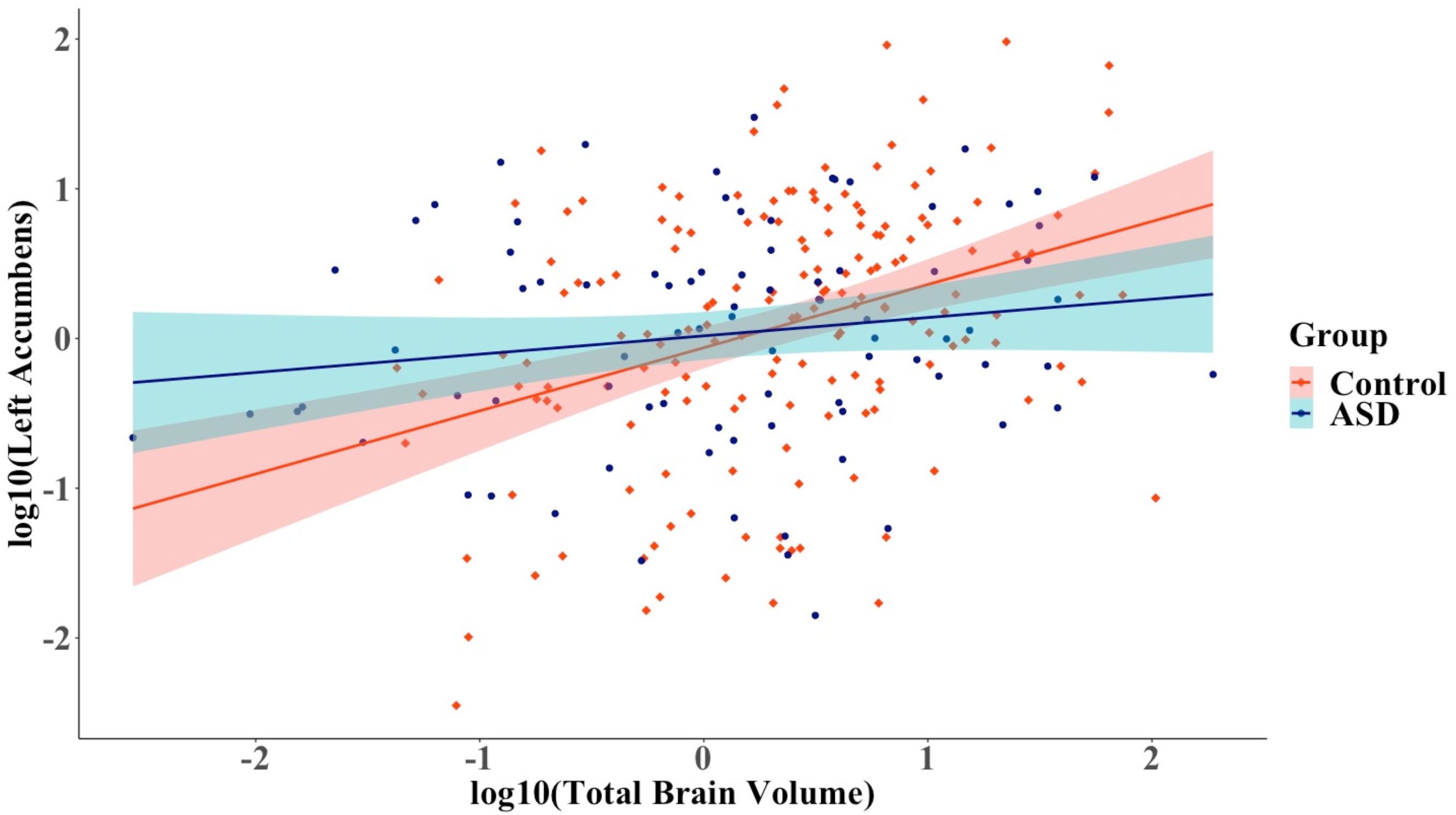
Relationship between the left accumbens and total brain volume across groups after outlier and comorbidity removal (NControl = 167, N ASD = 85) in boys with a Full Scale Intelligence Quotient < median (107.8). ASD, Autism Spectrum Disorder. 95% confidence region are given by group. Volumes were log transformed and scaled.

**Table 5.** Left Accumbens LMEM Results (A) and Unstandardized Allometric Coefficients (B) for Boys with a Full Scale Intelligence Quotient > median (107.8)

| A) | **Log10( Left Accumbens) ~ Group x Log10(TBV)** |  |  |  |  |  |
| --- | --- | --- | --- | --- | --- | --- |
|  | (N<sub>ASD</sub> = 100<br>& N<sub>C</sub> = 174) |  |  | Without Outliers & Comorbidities<br>(N<sub>ASD</sub> = 85 & N<sub>C</sub> = 167) |  |  |
| | $\beta$ | SE | pFDR | $\beta$ | SE | pFDR |
| Medication |  |  |  | 0.11 | 0.15 | 0.46 |
| Log10(TBV) | 0.54 | 0.08 | $2.90 \times 10^{-11}$ | 0.41 | 0.07 | $2.92 \times 10^{-8}$ |
| Group | 0.12 | 0.10 | 0.295 | 0.02 | 0.10 | 0.822 |
| Group x<br>log10(TBV) | -0.32 | 0.10 | 0.003 | -0.24 | 0.10 | 0.044 |

| B) | **Log10( Left Accumbens) ~ Group x Log10(TBV)** |  |  |  |  |  |
| --- | --- | --- | --- | --- | --- | --- |
|  | (N<sub>ASD</sub> = 100<br>& N<sub>C</sub> = 174) |  |  | Without Outliers & Comorbidities<br>(N<sub>ASD</sub> = 85 & N<sub>C</sub> = 167) |  |  |
| | $\alpha$ | SE | pFDR | $\alpha$ | SE | pFDR |
| log10(TBV) |  |  |  |  |  |  |
| ASD (N = 100) | 0.45 | 0.15 | 0.009 | 0.26 | 0.18 | 0.377 |
| Control (N = 174) | 1.21 | 0.18 | 0.000 | 0.97 | 0.16 | 0.000 |

*N.B.* $\beta$ corresponds to standardized beta for all main effects and interactions. $\alpha$ corresponds to the unstandardized allometric scaling coefficient of log10(TBV) with log10(left Accumbens). C corresponds to controls, FDR to false discovery rate correction for multiple comparison, and TBV to Total Brain Volume.

Although the left pallidum had a group difference over 0.2 (ß > 0.2) in the MGCFA, which was replicated in the corresponding LMEM after FDR correction (ß = -0.26, SE = 0.10, p = 0.023), the allometric scaling group difference was no longer significance after including medication as a covariate and removing outliers and comorbidities (ß = 0.08, SE = 0.11, p = 0.607). Although cortical white matter was not investigated in the MGCFA due to model convergence issues, allometric scaling did not differ between groups (ß = 0.00, SE = 0.04, p = 0.864).

To examine if the allometric scaling group difference reported in the left accumbens of boys with an FSIQ > median depended on age, we ran a LMEM with the left accumbens as dependent variable, TBV by group by Age (linear or quadratic) as fixed effects and scanner site as random intercept (Table 6.A). Again, ASD individuals had a smaller allometric scaling coefficient compared to controls before and after outlier and comorbidity removal and medication use inclusion (Table 6.A & B). Linear age and age effects were similar, although the effect sizes were slightly greater in the model with quadratic age (Table S18).

Post hoc analyses revealed that the total ADOS did not significantly predict left accumbens volume (ß = - 0.01, SE = 0.02, p = 0.770) or the allometric scaling relationship (ß = -0.02, SE = 0.02, p = 0.770) of that volume in ASD individuals with an available total ADOS score (N = 59).

**Table 6.** Left Accumbens LMEM Results with Age (A) and Unstandardized Allometric Coefficients (B) for Boys with a Full Scale Intelligence Quotient > median (107.8)

| A) | **Log10( Left Accumbens) ~ Group x Log10(TBV) x Age** |  |  |  |  |  |
| --- | --- | --- | --- | --- | --- | --- |
|  | (N<sub>ASD</sub> = 100<br>& N<sub>C</sub> = 174) |  |  | Without Outliers &<br>Comorbidities<br>(N<sub>ASD</sub> = 79 & N<sub>C</sub> = 162) |  |  |
| | $\beta$ | SE | pFDR | $\beta$ | SE | pFDR |
| Medication |  |  |  | 0.15 | 0.15 | 0.551 |
| Log10(TBV) | 0.55 | 0.07 | $1.89 \times 10^{-11}$ | 0.48 | 0.07 | $6.63 \times 10^{-10}$ |
| Group | 0.07 | 0.10 | 0.618 | 0.03 | 0.10 | 0.804 |
| Age | -0.14 | 0.07 | 0.12 | -0.12 | 0.06 | 0.132 |
| Log10(TBV) x Group | -0.28 | 0.10 | 0.024 | -0.32 | 0.10 | 0.011 |
| Log10(TBV) x Age | 0.04 | 0.07 | 0.705 | 0.04 | 0.07 | 0.783 |
| Group x Age | 0.04 | 0.10 | 0.705 | -0.02 | 0.09 | 0.804 |
| Group x log10(TBV) x Age | 0.15 | 0.10 | 0.311 | 0.21 | 0.12 | 0.176 |

B)
|  | **Log10( Left Accumbens) ~ Group x Log10(TBV) x Age** |  |  |  |  |  |
| --- | --- | --- | --- | --- | --- | --- |
|  | (N<sub>ASD</sub> = 100<br>& N<sub>C</sub> = 174) |  |  | Without Outliers & Comorbidities<br>(N<sub>ASD</sub> = 79 & N<sub>C</sub> = 162) |  |  |
| log10(TBV) | $\alpha$ | SE | pFDR | $\alpha$ | SE | pFDR |
| ASD (N = 100) | 0.54 | 0.16 | 0.005 | 0.27 | 0.19 | 0.280 |
| Control (N = 174) | 1.26 | 0.18 | 0.000 | 1.10 | 0.16 | 0.000 |

*N.B.* $\beta$ corresponds to standardized beta for all main effects and interactions. $\alpha$ corresponds to the unstandardized allometric scaling coefficient of log10(TBV) with log10(left Accumbens). C corresponds to controls, FDR to false discovery rate correction for multiple comparison, and TBV to Total Brain Volume.

#### Global Volumetric Group Differences

Scalar invariance was supported in boys from 6 to under 12 years old (∆χ2_(22)_ = 24.37, p = 0.328), in boys aged 12 to under 20 years old (∆χ2_(22)_ = 30.6, p = 0.104), boys with an FSIQ ≤ 107.8 (∆χ2_(22)_ = 28.0, p = 0.176), and in boys with an FSIQ >107.8 (∆χ2_(22)_ = 27.5, p = 0.194), suggesting that there are no volumetric differences between ASD and TD individuals when correcting for individual differences in TBV by taking into account allometric scaling. However, unlike the MGCFA, LMEMs in the right hippocampus of boys from 12 to under 20 years old revealed a group difference in volume adjusted for allometric scaling (Table 4). Specifically, ASD individuals (M = 4300.41 mm^3^, SD = 501.38 mm^3^) had a greater volume than their control counterparts (M = 4201.98 mm^3^, SD = 582.45 mm^3^). A volumetric difference in the left caudate was also found for boys with an FSIQ > the median before (ß = 0.30, SE = 0.12, p = 0.013) and after including medication as a covariate and removing comorbidities and outliers (ß = 0.30, SE = 0.10, p = 0.011; *a posteriori* Power Analyses Table *S17*). Specifically, ASD individuals (M = 4299.23 mm^3^, SD = 451.55 mm^3^) had a greater volume their control counterparts (M = 4227.20 mm^3^, SD = 646.23 mm^3^).

### IV- Comparing TBV Adjustment Techniques for Individual Differences in TBV

#### Present Study

For the right hippocampus in the sample of boys from 12 to under 20 years old, the linear covariate and allometric scaling TBV adjustment technique revealed volumetric group differences that were absent when omitting TBV and applying the linear interaction adjustment (Table 7.A). However, TBV adjustment techniques yielded similar results for the left accumbens in the sample of boys with and FSIQ > median (107.8; Table 7 B.). Overall, these results suggest that the extent to which the type of adjustment techniques influences reported volumetric and scaling group differences varies across GM volumes.

**Table 7.** Variations in Exploratory Neuroanatomical Group Differences across TBV Adjustment Techniques without outliers and comorbidities.

A)
| **Right Hippocampus** | Effect | *B* | SE | pFDR |
| --- | --- | --- | --- | --- |
| *No Adjustment*<br>Group x FSIQ + Medication | Group | 0.25 | 0.13 | 0.083 |
| *Linear Covariate Adjustment*<br>TBV + Group x FSIQ + Medication | Group | 0.27 | 0.10 | 0.027 * |
| *Linear Interactive Adjustment*<br>TBV x Group x FSIQ + Medication | Group | 0.26 | 0.10 | 0.054 |
|  | Group by TBV | -0.21 | 0.10 | 0.103 |
| *Allometric Interactive Adjustment*<br>Log10(TBV) x Group x FSIQ + Medication | Group | 0.21 | 0.07 | 0.021 * |
|  | Group by log10(TBV) | -0.22 | 0.09 | 0.037 * |

B)
| **Left Accumbens** | Effect | *B* | SE | pFDR |
| --- | --- | --- | --- | --- |
| *No Adjustment*<br>Group x Age + Medication | Group | -0.08 | 0.13 | 0.948 |
| *Linear Covariate Adjustment*<br>TBV + Group x Age + Medication | Group | -0.06 | 0.12 | 0.859 |
| *Linear Interactive Adjustment*<br>TBV x Group x Age + Medication | Group | -0.11 | 0.12 | 0.546 |
|  | Group by TBV | -0.29 | 0.10 | 0.022 * |
| *Allometric Interactive Adjustment*<br>Log10(TBV) x Group x Age + Medication | Group | 0.03 | 0.10 | 0.804 |
|  | Group by log10(TBV) | -0.32 | 0.10 | 0.011 * |

*N.B.* $\beta$ corresponds to standardized beta, TBV to total brain volume, FSIQ to Full Scale Intelligence Quotient, and FDR to False Discovery Rate correction for multiple comparisons (\* : significance at 0.05 after FDR correction).

#### Replication of Zhang and colleagues (2018)

In the LMEMs without TBV adjustment, we replicated the significant interaction of group by linear age by sex in the hippocampus. We were unable to replicate the remaining group differences reported by Zhang and colleagues (2018; Table 8). Although Zhang and colleagues (2018) reported that the interaction of group by linear age by sex in the hippocampus was no longer significant when covarying for TBV (no statistics were provided), the interaction remained minimally significant in our sample (Table 8).

**Table 8.**
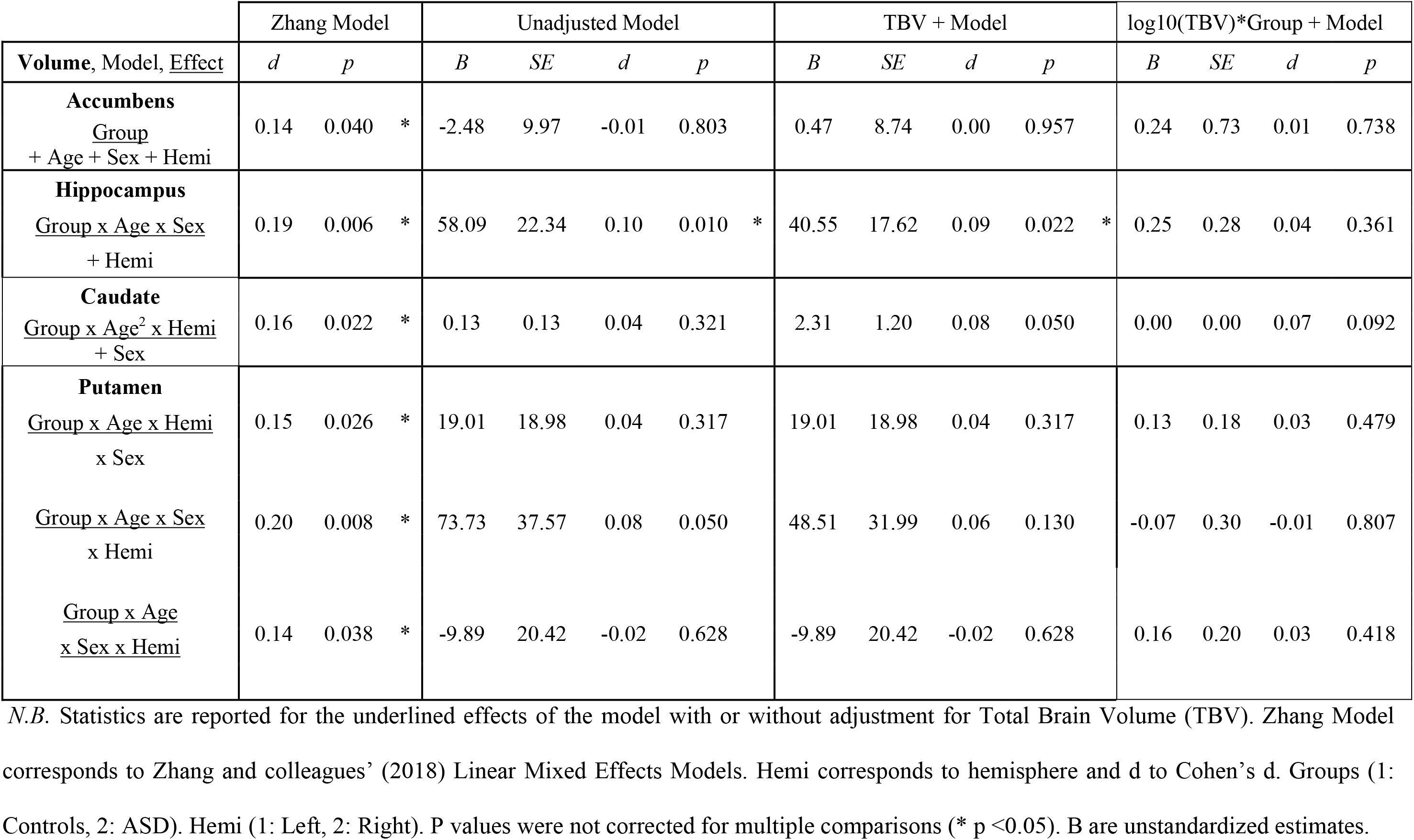
Replication of the Significant Group Effects Reported by Zhang and colleagues (2018) and TBV Adjustment Comparison.

When comparing results from LMEM across all brain volumes with varying TBV adjustment techniques (Table 8 and Tables S19 – S27), we found that the effect size of TBV was smaller when considering allometric scaling across all volumes. Although generally consistent, there were some differences in effect size and significance across TBV adjustment techniques. For instance, the interaction of group by linear age by sex in the hippocampus previously reported in LMEMs without TBV and with linear TBV adjustment was no longer significant when adjusting for TBV with allometric scaling (Table 8). Instead, the interaction of group by log10 (TBV) by sex was significant (ß = -0.40, SE = 0.20, p = 0.041, d = -0.08) when linear age was included in the model (Table S19). The interaction was no longer significant following FDR correction for multiple comparisons and was not significant when linear age was included in the model.

## Discussion

The primary aim of this study was to investigate subcortical allometric scaling and volumetric differences between TD and ASD individuals from the ABIDE I while adjusting for individual differences in TBV by taking into account brain allometry. The secondary goal of this paper was to identify if subcortical allometric scaling and volumetric group differences depended on sex, age, and/or FSIQ. We compared the results of two statistical methods: MGCFAs, which advantageously test global and regional cerebral group differences while considering the mutual relationships between volumes, and LMEMs, to evaluate result consistency across methods and facilitate result comparison with the literature on volumetric differences in ASD. MGCFAs and LMEMs were generally consistent. While no robust neuroanatomical group differences were reported in the entire sample, exploratory MGCFAs and LMEMs revealed group differences in allometry for the right hippocampus in boys aged 12 to under 20 years old and the left accumbens in boys with an FSIQ > median. In terms of adjustment techniques for TBV, our findings further support that the type of adjustment techniques for TBV can influence reported volumetric and scaling group differences and suggest that allometric scaling should be considered to reduce the risk of reporting biased neuroanatomical group differences.

### Allometric Scaling in ABIDE

While the right and left cortex were isometric (α = 1) and cerebral white matter was hyperallometric (α > 1), the majority of subcortical volumes in TD and ASD individuals were hypoallometric (α < 1). Scaling coefficients in controls were generally consistent with the few available in the literature (Liu et al., 2014; Reardon et al., 2016). Following outlier removal, the scaling coefficient of the left accumbens and right amygdala in controls were also isometric when sex and age effects were considered. While our findings could suggest that allometry is not a characteristic of all brain regions, a recent study examining surface area scaling coefficients reported different scaling coefficients within brain regions (e.g. both, negative and positive scaling in the amygdala (Reardon et al., 2018). Brain allometry should in turn be investigated in cortical and subcortical subregions (not examined in the present study) since allometric scaling across these regions may serve as cerebral markers of ASD.

### Group Differences in TBV

TBV only differed between ASD and TD individuals in the sample of boys with an FSIQ ≤ 107.8. TBV was greater for individuals with ASD compared to their control counterparts. However, this difference in TBV between groups may be artefactual considering that IQ and brain size are differently correlated between ASD subjects (r = 0.08) and controls (r = 0.31). The study which provided us the ABIDE I data simulated the impact of matching patient and control subjects by FSIQ and reported that FSIQ matching can bias TBV group differences by increasing the number of patient with a large TBV (Lefebvre et al., 2015). The biasing effect of IQ matching on TBV differences may also explain why one ABIDE study reported a subtle TBV group differences (1-2%) after controlling for IQ in the matched but not the entire cohort (Riddle et al., 2017).

The lack of a general TBV difference is consistent with past ABIDE studies examining volumetric group differences (Haar et al., 2016; Riddle et al., 2017; Zhang et al., 2018). While previous studies reported neuroanatomical differences between ASD and TD individuals across stages of development (Duerden et al., 2012; Stanfield et al., 2008), no group differences in TBV were found in children and adolescent boys in the present study. Since the studies that report a greater TBV in children with ASD suggest that TBV group differences are greater in early childhood and are no longer apparent in 10 year old children (Courchesne et al., 2011; Lange et al., 2015), the absence of TBV group differences in the investigated children may be due to that fact that the children in our subsample were too old to exhibit TBV group differences (1^st^ Quartile = 9.3 years old). In terms of studies reporting TBV group differences in adolescents, the majority of studies were either underpowered (Freitag et al., 2009; Hazlett et al., 2005) or regrouped both adolescent and children (Duerden et al., 2012), suggesting that their findings may be unreliable or biased by the younger children included in their sample. The present study provides further evidence that enlarged TBV may not serve as a reliable biomarker of ASD after young childhood and may instead represent a bias in the population norm (Raznahan et al., 2013).

### No Regional Volume Group Differences in the Entire Sample

ASD and TD individuals did not differ in terms of the presently investigated cortical and subcortical volumes. Although consistent with recent large scale studies (Riddle et al., 2017; Zhang et al., 2018), this finding contrasts with the largest study to our knowledge (N _Asd_ = 1, 571 & N _Controls_ 1, 651; van Rooij et al., 2017) examining cortical and subcortical differences in ASD. The authors linearly adjusted for TBV (covariate approach) and reported volumetric group differences in the pallidum, putamen, amygdala, and nucleus accumbens (Cohen’s d = -0.08 to -0.13). While the absence of such small volumetric group differences may stem from our smaller sample size, the covariate approach for TBV adjustment has also been shown to yield a higher rate of false positives (Liu et al., 2014; Sanchis-Segura et al., 2019), suggesting that these results should be replicated with an allometric scaling adjustment for TBV to be judged robust.

Volumetric group differences may lie in other cortical areas and WM volumes that make up the large-scale neurocognitive systems assumed to mediate symptoms of ASD. However, reported group differences in cortical regions (e.g. the insula (Radeloff et al., 2014) and prefrontal cortex (Duerden et al., 2012) thought to be involved in social cognition (Blakemore, 2008)) and in WM volumes (e.g. corpus callosum assumed to enable the integration of multiple sources of stimulation; Just et al., 2007) must be replicated in sufficiently powered studies (Di & Biswal, 2016; Haar et al., 2016; Lefebvre et al., 2015) that appropriately adjust for TBV (Liu et al., 2014; Sanchis-Segura et al., 2019) to be judged as robust neuroanatomical markers of ASD.

### No Regional Volume Age, Sex, & FSIQ Dependent Group Differences in the Entire Sample

The age and sex dependent volumetric group differences reported by the ABIDE I study we aimed to replicate (Zhang et al., 2018) and by several cross-sectional studies and meta-analyses on the neuroanatomical variations of ASD (Duerden et al., 2012; Greimel et al., 2013; D. Y.-J. Yang et al., 2016; X. Yang et al., 2016) were not replicated in this study. These discrepancies with the literature may stem from (i) limited statistical power, (ii) publication bias in favor of positive results and (iii) from the lack of correction for multiple comparison across a majority of studies, which increases the risk of false positives. In fact, consistent with our entire sample analyses, the largest-scale ASD study to date that addresses these limitations did not report age by sex or age by diagnostic effects when the linear effects of age were considered (van Rooij et al., 2017). However, based on previous findings that omitting brain allometry can lead to underestimating group differences (Mankiw et al., 2017; Reardon et al., 2016), we cannot rule out the presence of small age by sex or age by diagnostic effects on the investigated regional volumes since they would not be detectable with our current sample size.

Unlike the largest study to date on cerebral markers of ASD, which linearly corrected for TBV (covariate approach) and found volumetric sex differences in the thalamus, caudate, putamen, amygdala, and nucleus (van Rooij et al., 2017), no sex effects were found in our study. Although the absence of sex effects may be due to the few females (N = 106) in our sample, some significant sex effects may be false positives considering that the covariate TBV adjustment tends to overestimate volumetric sex differences (Reardon et al., 2016; Sanchis-Segura et al., 2019). In light of the numerous methodological discrepancies in the studies on the neuroanatomical group differences in ASD, more large scale studies with an allometric scaling adjustment for TBV will be necessary to unbiasedly estimate cerebral differences in ASD across sexes.

### Regional Volume Age, Sex, & FSIQ Dependent Group Differences in Exploratory Subsamples

Based on the LMEMs in the entire sample, allometric scaling and volumetric group differences did not depend on sex, age, and/or FSIQ. Exploratory analyses were nonetheless run on subsamples previously examined in the literature on the subcortical correlates of ASD (e.g. Lin et al., 2015; Maier et al., 2015) to compare our findings with previous studies and to further examine result consistency between the MGCFA and LMEMs. Exploratory MGCFAs and LMEMs revealed that allometric scaling coefficients were smaller for ASD compared to controls in the right hippocampus for boys aged 12 to under 20 years old and in the left accumbens for boys with an FSIQ < median. This finding suggests that although both groups had hypoallometric scaling coefficients, indicating that these regional volumes grow at a slower rate than TBV, regional volume increased less with TBV in ASD individuals compared to controls.

Hypoallometry (exponent <1) in the right hippocampus and left accumbens regions of ASD boy subsamples did not covary with ASD severity. Yet, previous studies reported that the neuroanatomy of ASD is heterogeneous and varies with ASD severity (Bedford et al., 2020; H. Chen et al., 2019). One possibility is that the size of the present sample may not be sufficient to report a link between the allometric scaling coefficient and ASD severity, suggesting the presence of neuroanatomical differences between individuals with ASD based on ASD severity. On the other hand, it could also be that the severity of ASD is not linked to allometry or to subcortical structures which were presently investigated.

While allometric scaling group differences were consistent across methods, LMEMs revealed a greater right hippocampal volume in boys from 12 to under 20 which was not present in the MGCFA. Discrepancies in how parameter values are estimated in LMEMs and MGCFAs may explain inconsistencies across methods. For instance, unlike the LMEMs, the MGCFA (i) considers all regional volumes when predicting allometric scaling and volumetric group differences and (ii) takes into account correlated residuals when estimating parameter values. Yet, in light of the absence of allometric and volumetric group differences when examining the entire sample and the exploratory nature of these results, we recommend that these exploratory results be replicated in a larger sample to be judged robust.

### MGCFAs & LMEMs: Methodology

Although MGCFA and LMEMs generally provided similar results, we suggest that MGCFAs may not be optimal to investigate neuroanatomical differences between groups in future studies for several reasons. First, although the MGCFA can simultaneously conduct global and regional tests, the MGCFA cannot simultaneously examine FSIQ, age, and sex effects, factors thought to influence brain anatomy (Duerden et al., 2012; Mankiw et al., 2017; Reardon et al., 2016; Sacco et al., 2015; van Rooij et al., 2017; Zhang et al., 2018). However, considering that the primary goal was to examine neuroanatomical group differences regardless of age, sex, and FSIQ, the use of the MGCFA was appropriate. Second, the latent construct in the MGCFA cannot be equated with log10(TBV) which is typically employed to examine allometric scaling (Finlay et al., 2001), as in the LMEMs. Instead, the latent construct reflects the shared variance between the observed variables: the log-transformed regional volumes. Third, numerous correlated residuals (overlap in variance between volumes that measure something else than TBV) were included in each MGCFA to reach appropriate fit and these correlated residuals slightly differed in the entire sample and each subsample. Since brain regions across and within hemispheres are highly interconnected, the measurement error of one volume likely correlates with the measurement error of another volume. However, it is unclear to what extent the correlated residuals established in the present model reflect general relationships between brain regions, and to what extent they reflect idiosyncratic properties of the present sample. Only a comparison with another large dataset would allow one to assess how generalizable this model is. Nonetheless, we emphasize that the model fit of all MGCFAs were similar across groups and the results between LMEMs and MGCFAs were overall consistent.

Fourth, while the number of participants included in each subsample was sufficient to provide a MGCFA factor solution in agreement with the population structure from which the sample was taken (Mundfrom et al., 2005), more MGCFA simulation studies and the development of packages to estimate MGCFA power are needed to establish the number of participants required to observe a specific group difference in parameter (slope or intercept) at 80% power. Finally, additional simulation studies are required to ensure that the current MGCFA thresholds employed in the literature reflect ‘real’ rather than mathematical differences (Putnick & Bornstein, 2016). In the present study, Chen’s (2007) cutoff values for fit indices to determine regional metric invariance between groups were too conservative to detect the small neuroanatomical group differences reported by the χ2 difference tests and the LMEMs. One possibility is that Chen’s (2007) cutoff values for fit indices may be appropriate for testing invariance between groups on medium effect sizes but not for testing the small differences in parameter values in the current paper. Yet, this interpretation requires validation from future simulations studies conducted to identify appropriate fit indices cutoff values to detect small group differences in parameters in models with a varying number of factors and observed variables.

### Replication of Zhang and colleagues (2018)

Although the present paper used similar inclusion/exclusion criteria and analyzed data from the same cohort with the same statistical procedure, the only robust reproducible result from the latest ABIDE I study was the significant interaction of group by age by sex in the hippocampus without a TBV adjustment. Discrepancies between our findings and Zhang and colleagues’ (2018) can be explained by several factors. First, the small effect size and borderline p-values of the interactions reported by Zhang and colleagues (2018), suggest that these interactions without correcting for multiple comparisons were weak and perhaps not reliable. Second, while we selected similar age and FSIQ inclusion criteria, segmentation and quality checks differed between studies. While Zhang and colleagues (2018) used the FMRIB’s Automated Segmentation Tool (FAST) from the FMRIB’s Software Library (FSL), the present study used FreeSurfer. As a result, the mean of the investigated regional volumes and the distribution of participants across scanner sites for each volume somewhat differed between studies. Third, the current study’s smaller sample size (N = 654) following segmentation and quality checks may explain why fewer significant interactions were found compared to Zhang and colleagues (2018; N = 859). Inconsistencies between the present and replicated study provide further evidence for the fragility of many reported results and emphasize the need to reexamine results and identify the reasons for failures in replication to improve future research (Button et al., 2013).

### Comparing TBV Adjustment Techniques

In line with previous findings (Barnes et al., 2010; Mankiw et al., 2017, 2017; Sanchis-Segura et al., 2019), neuroanatomical group differences depended on the techniques used to adjust for individual differences in TBV. Analyses from the replication revealed the volumetric group differences in the hippocampus identified without TBV and with linear TBV adjustment were no longer significant when adjusting for TBV with allometric scaling (effect size was halved). The change in effect size suggests that omitting brain allometry can overestimate volumetric group differences even in the absence of TBV group differences. We additionally compared TBV adjustment techniques in the right hippocampus in boys from 12 to under 20 years old and in left accumbens for boys with an FSIQ over the median. Consistent with our findings from the replication, the type of adjustment technique and number of predictors included in the exploratory models influenced reported neuroanatomical differences in some volumes (i.e. in the right hippocampus and not the left accumbens). In light of our results and the literature reporting an effect of the TBV adjustment technique on reported neuroanatomical group differences (Liu et al., 2014; Sanchis-Segura et al., 2019), we stress that future studies consider brain allometry by using the allometric scaling TBV adjustment technique to provide unbiased estimates of the cerebral markers of ASD.

### Limitations

The current paper is limited in its capacity to study sex, age, and FSIQ effects on allometric scaling and volumetric group differences due to the insufficient number of girls, adults aged over 20, and individuals with an FSIQ < 70 in the ABIDE I sample. Further research on these populations is necessary to better understand ASD’s etiology for numerous reasons. For instance, while some females exhibit symptoms similar to males at an early age, high functioning females are thought to have more efficient coping strategies than males, specifically in the social domain (Dworzynski et al., 2012; Lai et al., 2015, 2017), which mask the severity of their ASD until later in adolescence or adulthood (Lai et al., 2015). By examining such individuals, who in part differ in terms of ASD symptomatology, future studies may shed a light on the neuroanatomical markers related to specific traits of ASD. In light of the cognitive changes associated with age-related brain volume alterations in the adult population (Scahill et al., 2003; Takao et al., 2012; Vinke et al., 2018), more adults in the young adult and older adult age ranges must be scanned and studied to accurately depict how age influences neuroanatomical differences reported in ASD. Finally, given that 1/3 of ASD individuals have an FSIQ < 70 (Christensen et al., 2016) and that they have a high within-group variability at the genomic level (Srivastava & Schwartz, 2014), neuroanatomical variations in these individuals likely depend on specific genetic components, warranting the investigation of cerebral differences with an imaging genetics approach in this population (Jack & Pelphrey, 2017).

Yet, considering the heterogeneity of symptoms experienced by autistic individuals (Jack & Pelphrey, 2017; McIntyre et al., 2017; Rao et al., 2008) and the diverse genetic contributions to ASD (Ramaswami & Geschwind, 2018), multimodal approaches (e.g. imaging genetics) should be applied across ASD individuals to better characterize ASD’s heterogeneity within etiologically dissimilar samples. Despite the need to inspect diverse ASD samples to fully understand the heterogeneity of ASD, investigating neuroanatomical group differences in ABIDE I is a primordial step to identifying robust allometric scaling and volumetric group differences in high functioning (FSIQ > 70) children and adolescents.

### Implications

Correcting for TBV with allometric scaling provides more accurate estimates of group differences in cerebral volumes and investigates whether allometric scaling could serve as a neuroanatomical marker for group differences in behavior and cognition. However, while numerous studies have proposed functional correlates to regional volume changes, the influence of allometric scaling on behavior and cognition remains unknown. For instance, while a reduced hippocampal volume has previously been linked to impaired episodic memory (Salmond et al., 2005; Williams et al., 2006) and a decrease in the left putamen volume to greater repetitive and stereotyped behavior in ASD (Cheung et al., 2010; Estes et al., 2011), atypical allometric scaling relationships may or may not translate to such cognitive and behavioral symptoms. Yet, before linking cerebral markers to variations in cognition and behavior, robust neuroanatomical markers that consider additional factors thought to influence cerebral diversity in the TD (e.g. sex, age) and in the ASD (e.g. minimally verbal subtype, IQ) population must be established.

There are numerous efforts aimed at identifying cerebral markers of ASD with brain imaging techniques for diagnosis purposes (e.g. Alvarez-Jimenez et al., 2020; Kong et al., 2019; Nielsen et al., 2013). However, our study along with the increasing literature reporting the absence of (Haar et al., 2016; Lefebvre et al., 2015) or very subtle (van Rooij et al., 2017) volumetric group differences, suggest that previous group differences in subcortical volumes are potentially false positives or that individual regions may not constitute useful cerebral markers to employ for the diagnosis of ASD. This is consistent with the emerging literature that focuses on training classification algorithms with numerous brain regions and various methods, such as resting state functional MRI, to generate a diagnostic tool for ASD (Heinsfeld et al., 2018; Plitt et al., 2015). Thus, from a clinical standpoint, our findings further support that the cerebral markers of ASD, which could be used for diagnosis, should not be restricted to a specific region in the brain.

Once robust cerebral markers that covary with cognitive abilities and disease severity are identified, mediation models can be conducted by future studies to uncover the diverse causal links of ASD that integrate genetic, environmental, cognitive, and behavioral information (Lai et al., 2013). These advances may enable the creation of more accurate ASD subgroups, offer more accurate diagnostic criteria, which are increasingly being used to automate diagnosis (H. Chen et al., 2019; Nielsen et al., 2013), as well as facilitate person-centered treatment by providing insights on ASD’s complex etiology.

## Conclusion

The primary goal of this study was to identify allometric scaling and volumetric differences between TD and ASD individuals when taking into account brain allometry and the second goal to examine whether cerebral group differences depended on age, sex, and/or FSIQ. We analyzed data from ABIDE I using a commonly used univariate approach, LMEM, and a multivariate approach part of structural equation modeling, MGCFA. No robust allometric and volumetric group differences were observed in the entire sample, although exploratory analyses on subsamples based on age, sex, and FSIQ suggested that allometric scaling and volume may depend on age, sex, and/or FSIQ. While the LMEMs and the MGCFA were generally consistent, we propose that LMEMs may be more efficient to examine neuroanatomical group differences in light of the encountered methodological MGCFA constraints (e.g. no interaction effects, convergence issues, correlated residuals inclusion). Additional LMEM analyses with different TBV adjustment techniques revealed that the effect sizes and significance of cerebral differences between TD and ASD individuals differed across TBV adjustment techniques.

In addition to being the first study to examine allometric scaling and volumetric differences between ASD and TD individuals in the presently investigated volumes, the study adds to the literature by offering reference scaling coefficients for future studies in both ASD and TD individuals and by comparing two statistical methods: the MGCFA and LMEM. Finally, in its difficulty to replicate a recent similar study, the paper contributes to the literature on the replication crisis and, through its comparison of TBV adjustment techniques, provides further support for the consideration of brain allometry to reduce reporting biased estimates of neuroanatomical group differences.

## Data Availability

The data that support the findings of this study are openly available in Subcortical-Allometry-in-Autism at http://doi.org/10.5281/zenodo.3592884.

## Acknowledgements

This work received support under the program “Investissements d’Avenir” launched by the French Government and implemented by ANR with the references ANR-17-EURE-0017 and ANR-10-IDEX-0001-02 PSL

## Conflict of interest

On behalf of all authors, the corresponding author states that there is no conflict of interest.

## Data Sharing and Data Accessibility

The data that support the findings of this study are openly available in “Subcortical-Allometry-in-Autism” at http://doi.org/10.5281/zenodo.3592884.

## Supplementary Materials

**Table of Contents**

**1. Demographics by Scanner Sites. 2**

**2. Power Analyses 17**

**3. MGCFA & LMEMs Assumptions 18**

**4. Replication of Zhang and Colleagues (2018): Methodology 23**

**5. MGCFA Results 2**5

**6. Replication Results 30**

### 1. Demographics by Scanner Sites

The distribution of participant brain volumes, age, sex, FSIQ, and number of participants somewhat differed between scanner sites and varied between the present study and Zhang and coleagues (2018; e.g. Carnegie Melon scanner site participants were not included in the present study). Considering that a number of participants were removed from the analyses following segmentation quality checks, we examined if the remaining ASD individuals were still matched by age, sex, FSIQ, and scanner sites to controls. A total of 302 ASD participants were matched by sex, age, sites, and FSIQ to 302 control participants (MatchIt R package; Ho, Imai, King, & Stuart, 2011) and the remaining 50 unmatched controls were included in the analyses to increase the sample size.

Participants did not differ in across sites in terms of handedness (χ^2^ _(12)_= 14.8, p = 0.206) although they differed in sex ratio (χ2 _(14)_= 43.9, p < 0.001), Full Scale Intelligence Quotient (FSIQ) scores (χ^2^ _(14)_= 34.5, p = 0.002), and age (χ^2^ _(14)_= 250.5, p < 0.001, d = 1.53). The distribution of all brain volumes, age, and FSIQ of Autism Spectrum Disorder (ASD) and control participants differed by scanner site following visual inspection and varied from the distribution of participants by scanner sites as shown by Figures S1-13. The figures correspond to the same figures included in Zhang and colleagues’s (2018) supplemental information to facilite sample comparison between our study and Zhang and colleagues (2018). Zhang and colleagues’ (2018) supplementary figures are available [here](https://www.cambridge.org/core/journals/psychological-medicine/article/revisiting-subcortical-brain-volume-correlates-of-autism-in-the-abide-dataset-effects-of-age-and-sex/CB66FFA7347DBE59C446BA66B1BA1A66#fndtn-supplementary-materials).

**Figures S1.**
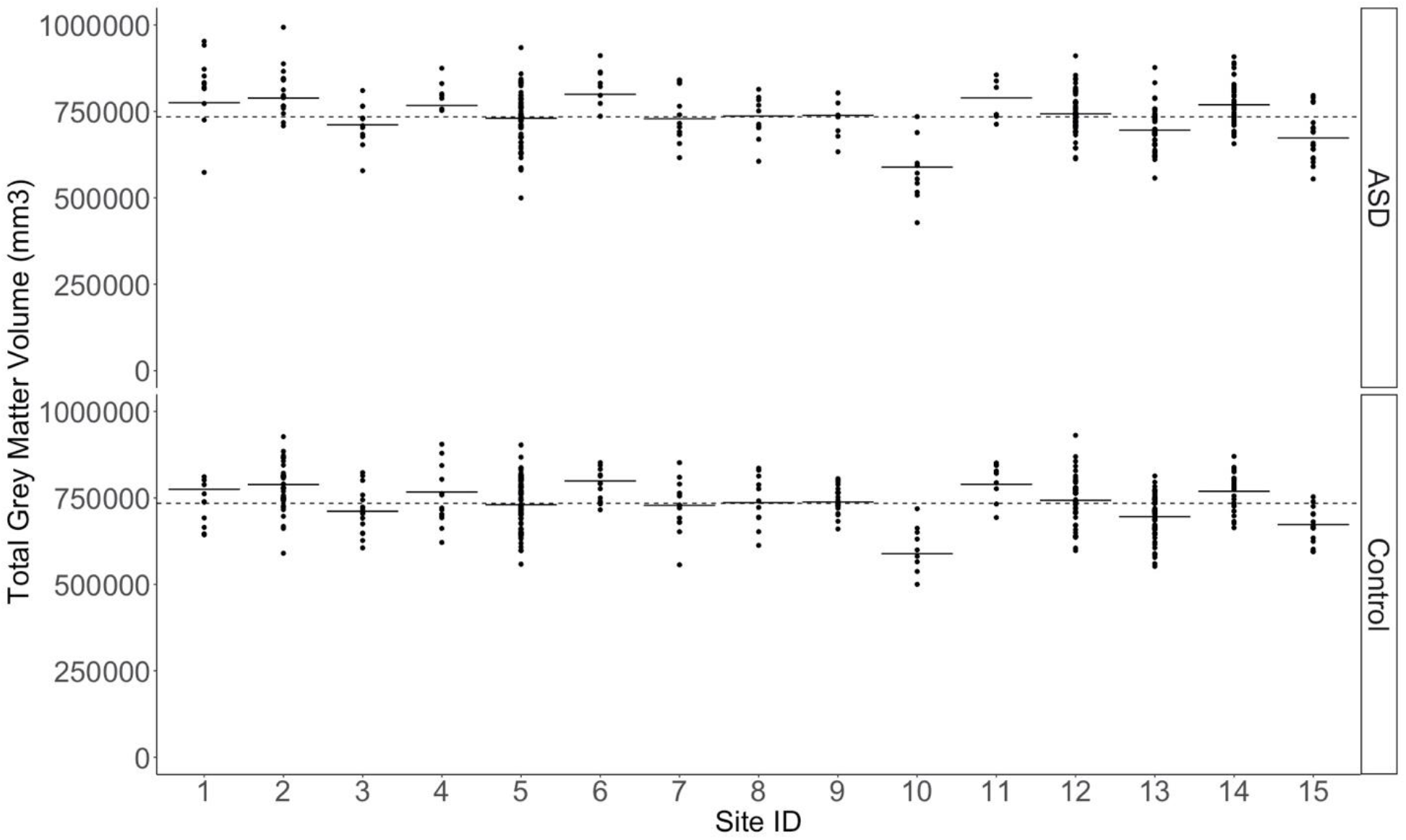
Distribution of Total Grey Matter Volumes of Autism Spectrum Disorder (ASD) and Control Participants by Scanner Site. Circles represent the brain volume of each participant. (1 = California Institute of Technology; 2 = Kenny Krieger Institute; 3 = University of Leuven Sample; 4= Ludwig Maximilians University Munich; 5 = NYU Langone Medical Center; 6 = Oregon Health and Science University; 7= Olin, Institute of Living at Hartford Hospital; 8= University of Pittsburgh School of Medicine; 9 = San Diego State University; 10 = Stanford University; 11 = Trinity Center for Health Sciences; 12 = University of California Los Angeles; 13 = University of Michigan Sample; 14 = University of Utah School of Medicine; 15 = Yale Child Study Center).

**Figures S2.**
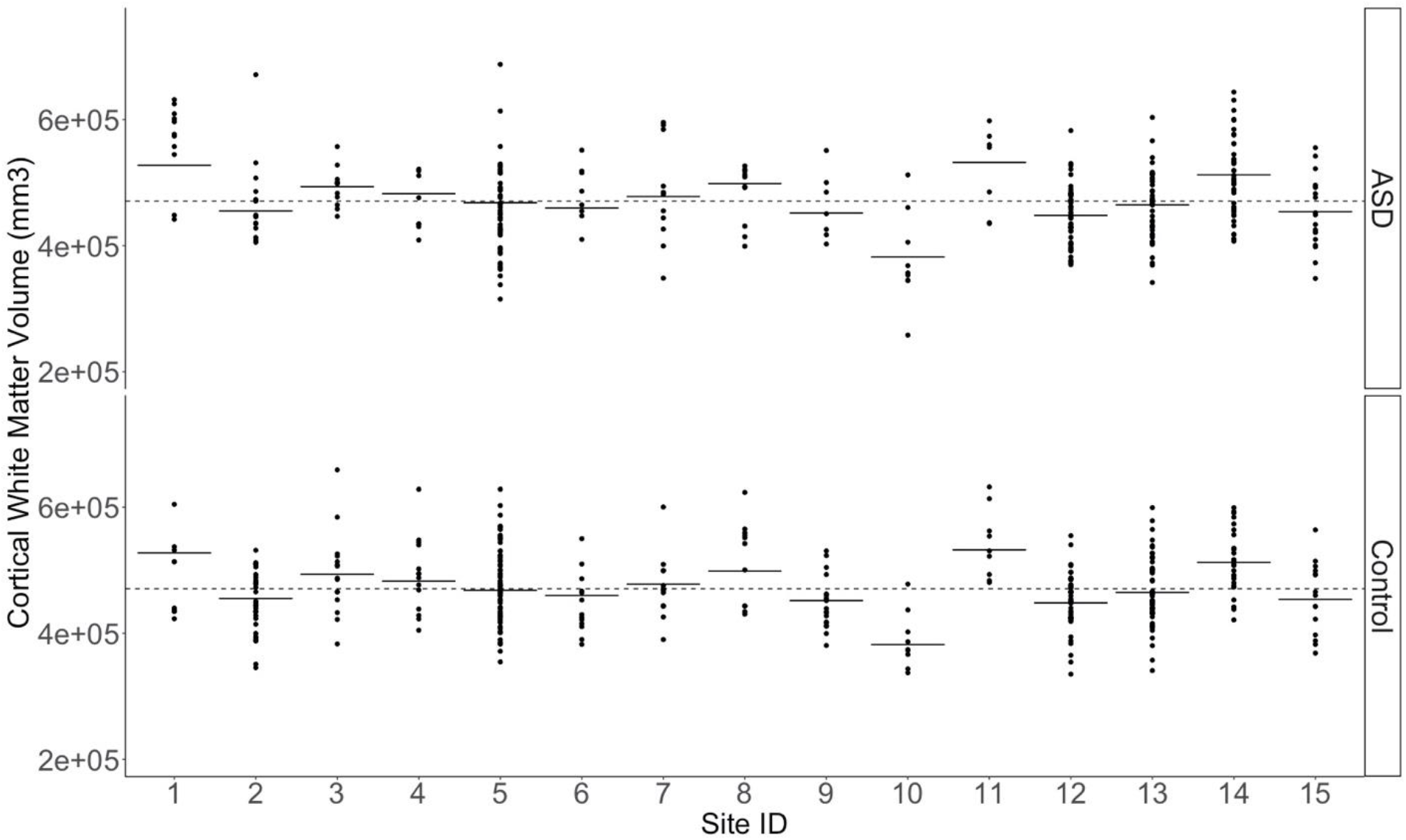
Distribution of Cortical White Matter Volumes of Autism Spectrum Disorder (ASD) and Control Participants by Scanner Site. Circles represent the brain volume of each participant. (1 = California Institute of Technology; 2 = Kenny Krieger Institute; 3 = University of Leuven Sample; 4= Ludwig Maximilians University Munich; 5 = NYU Langone Medical Center; 6 = Oregon Health and Science University; 7= Olin, Institute of Living at Hartford Hospital; 8= University of Pittsburgh School of Medicine; 9 = San Diego State University; 10 = Stanford University; 11 = Trinity Center for Health Sciences; 12 = University of California Los Angeles; 13 = University of Michigan Sample; 14 = University of Utah School of Medicine; 15 = Yale Child Study Center).

**Figures S3.**
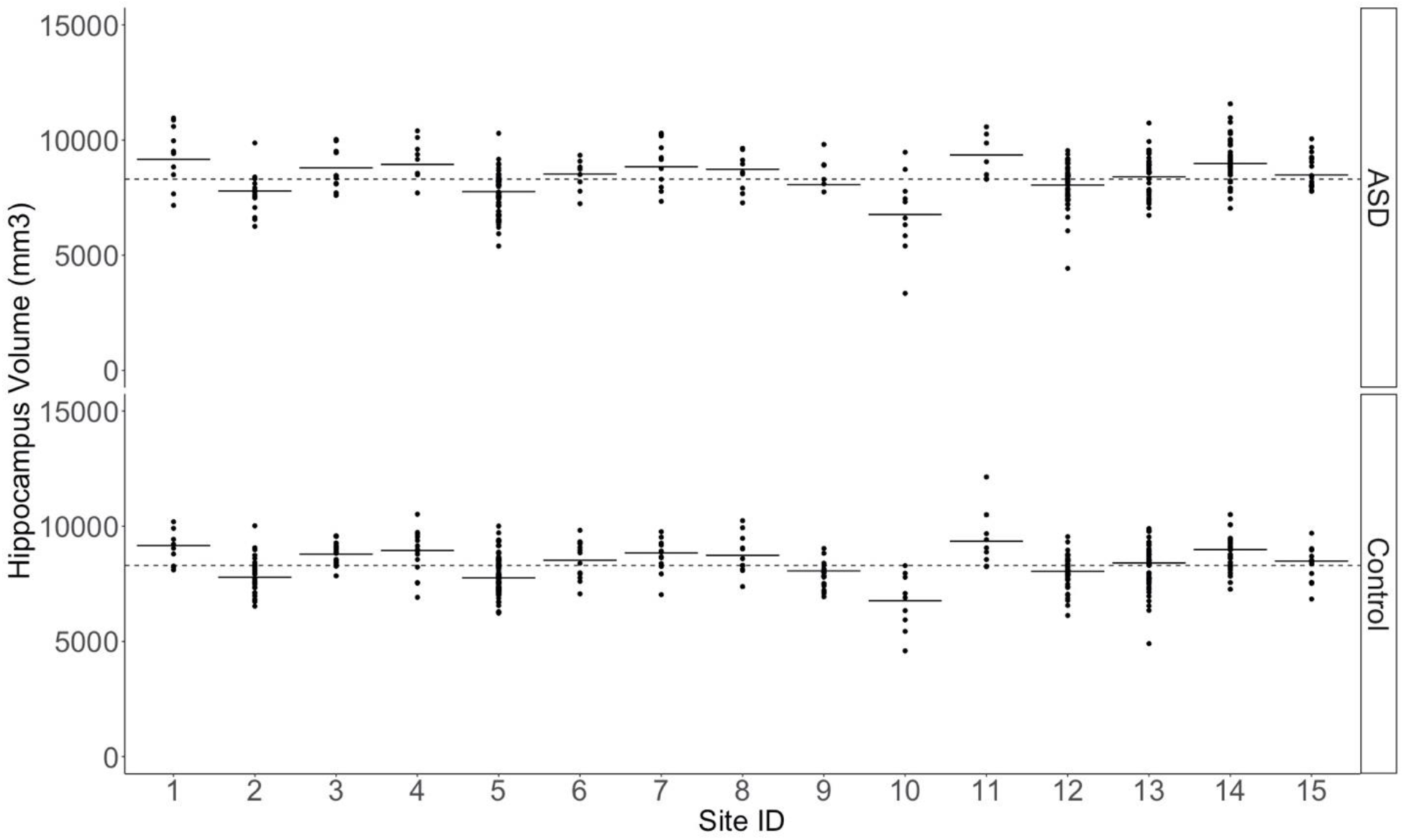
Distribution of Hippocampal Volumes of Autism Spectrum Disorder (ASD) and Control Participants by Scanner Site. Circles represent the brain volume of each participant. (1 = California Institute of Technology; 2 = Kenny Krieger Institute; 3 = University of Leuven Sample; 4= Ludwig Maximilians University Munich; 5 = NYU Langone Medical Center; 6 = Oregon Health and Science University; 7= Olin, Institute of Living at Hartford Hospital; 8= University of Pittsburgh School of Medicine; 9 = San Diego State University; 10 = Stanford University; 11 = Trinity Center for Health Sciences; 12 = University of California Los Angeles; 13 = University of Michigan Sample; 14 = University of Utah School of Medicine; 15 = Yale Child Study Center).

**Figures S4.**
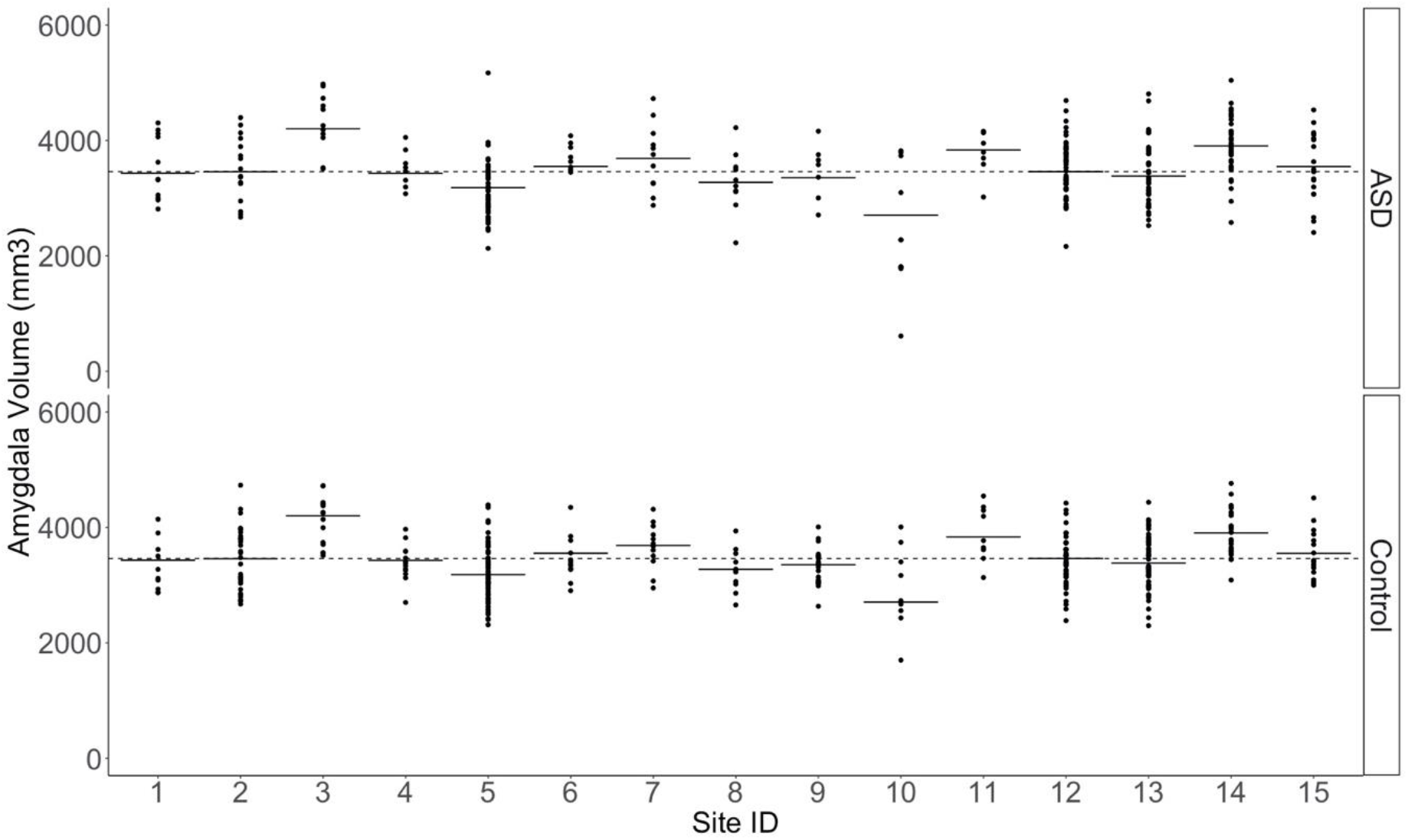
Distribution of the Amygdala Volumes of Autism Spectrum Disorder (ASD) and Control Participants by Scanner Site. Circles represent the brain volume of each participant. (1 = California Institute of Technology; 2 = Kenny Krieger Institute; 3 = University of Leuven Sample; 4= Ludwig Maximilians University Munich; 5 = NYU Langone Medical Center; 6 = Oregon Health and Science University; 7= Olin, Institute of Living at Hartford Hospital; 8= University of Pittsburgh School of Medicine; 9 = San Diego State University; 10 = Stanford University; 11 = Trinity Center for Health Sciences; 12 = University of California Los Angeles; 13 = University of Michigan Sample; 14 = University of Utah School of Medicine; 15 = Yale Child Study Center).

**Figures S5.**
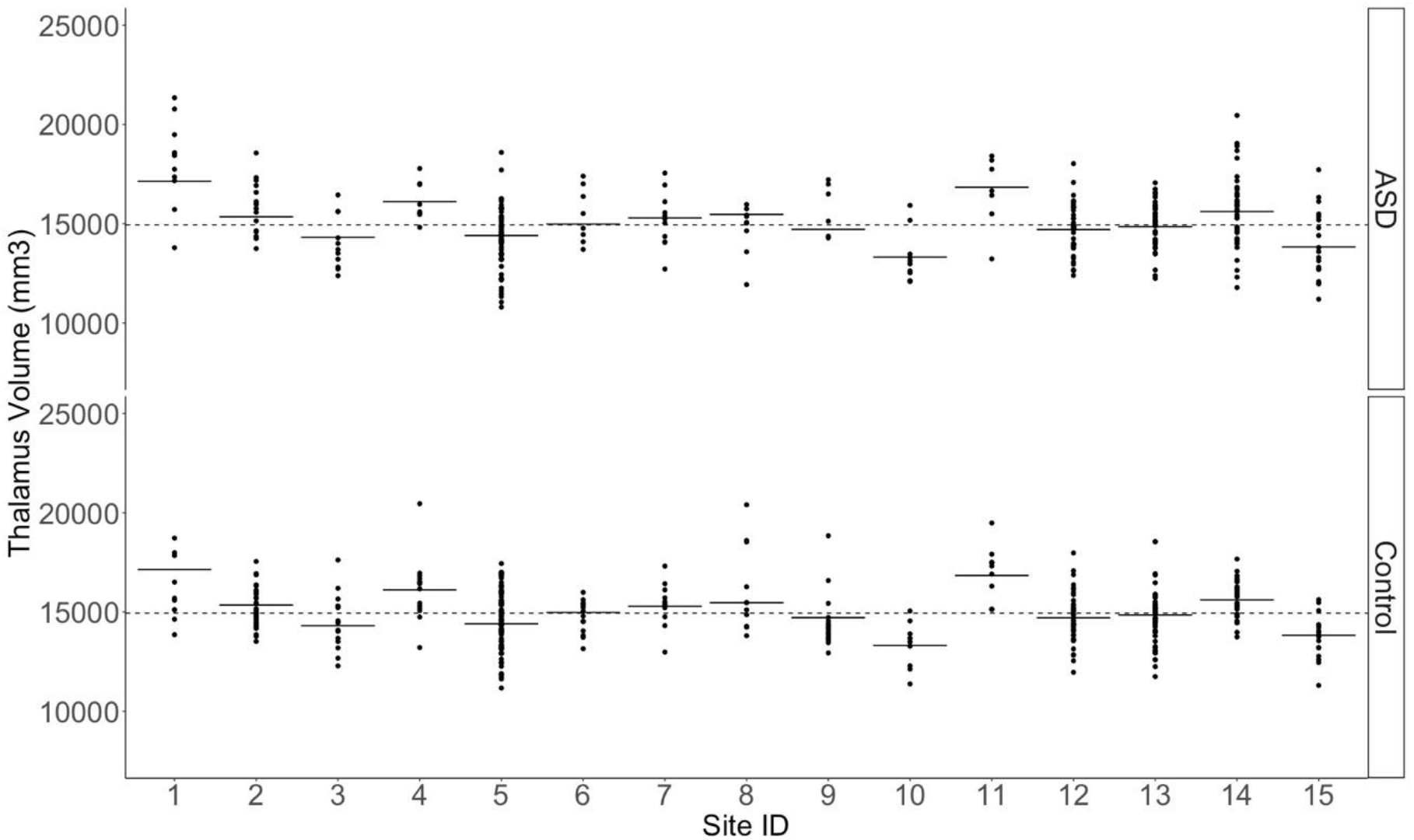
Distribution of Thalamic Volumes of Autism Spectrum Disorder (ASD) and Control Participants by Scanner Site. Circles represent the brain volume of each participant. (1 = California Institute of Technology; 2 = Kenny Krieger Institute; 3 = University of Leuven Sample; 4= Ludwig Maximilians University Munich; 5 = NYU Langone Medical Center; 6 = Oregon Health and Science University; 7= Olin, Institute of Living at Hartford Hospital; 8= University of Pittsburgh School of Medicine; 9 = San Diego State University; 10 = Stanford University; 11 = Trinity Center for Health Sciences; 12 = University of California Los Angeles; 13 = University of Michigan Sample; 14 = University of Utah School of Medicine; 15 = Yale Child Study Center).

**Figures S6.**
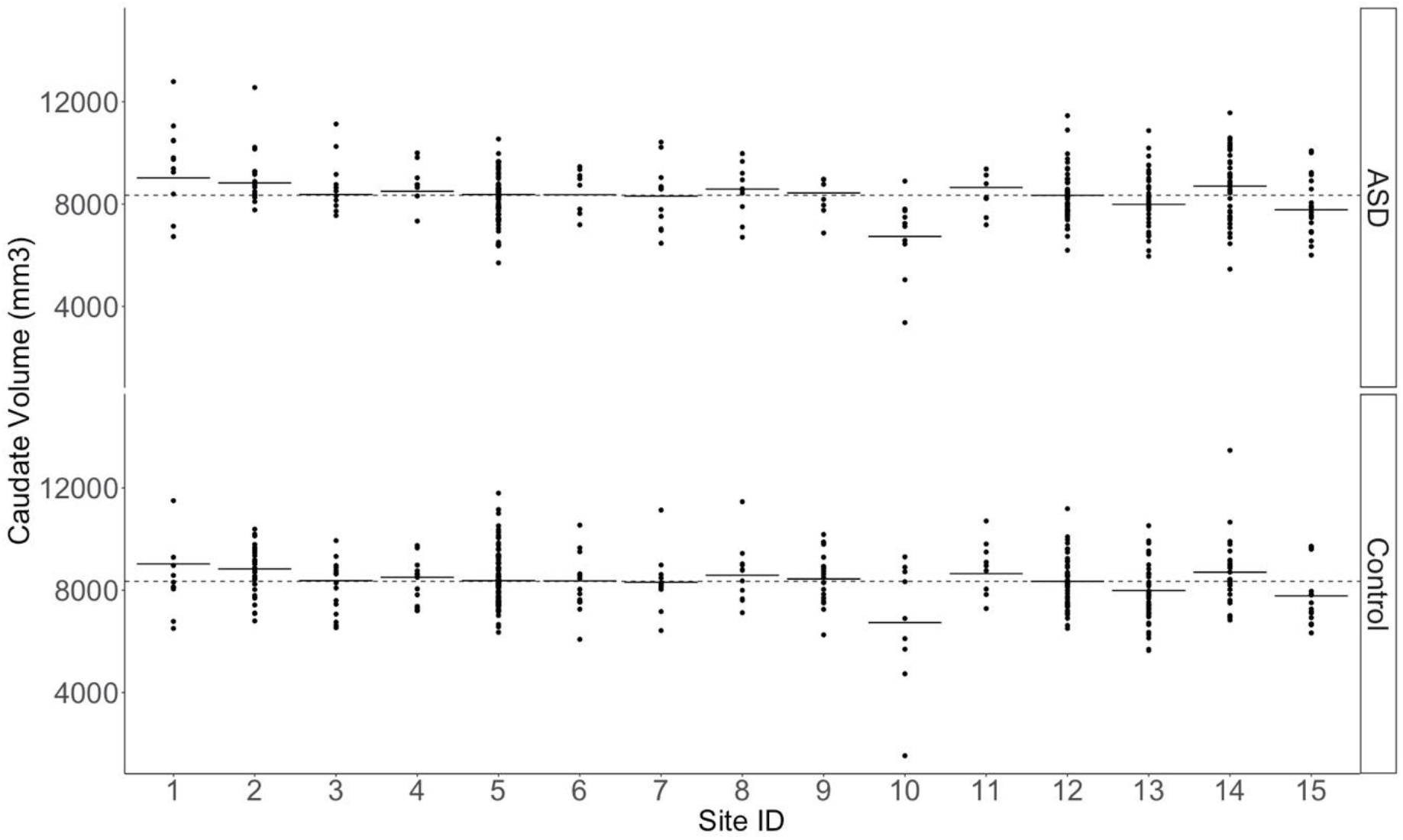
Distribution of Caudate Volumes of Autism Spectrum Disorder (ASD) and Control Participants by Scanner Site. Circles represent the brain volume of each participant. (1 = California Institute of Technology; 2 = Kenny Krieger Institute; 3 = University of Leuven Sample; 4= Ludwig Maximilians University Munich; 5 = NYU Langone Medical Center; 6 = Oregon Health and Science University; 7= Olin, Institute of Living at Hartford Hospital; 8= University of Pittsburgh School of Medicine; 9 = San Diego State University; 10 = Stanford University; 11 = Trinity Center for Health Sciences; 12 = University of California Los Angeles; 13 = University of Michigan Sample; 14 = University of Utah School of Medicine; 15 = Yale Child Study Center).

**Figures S7.**
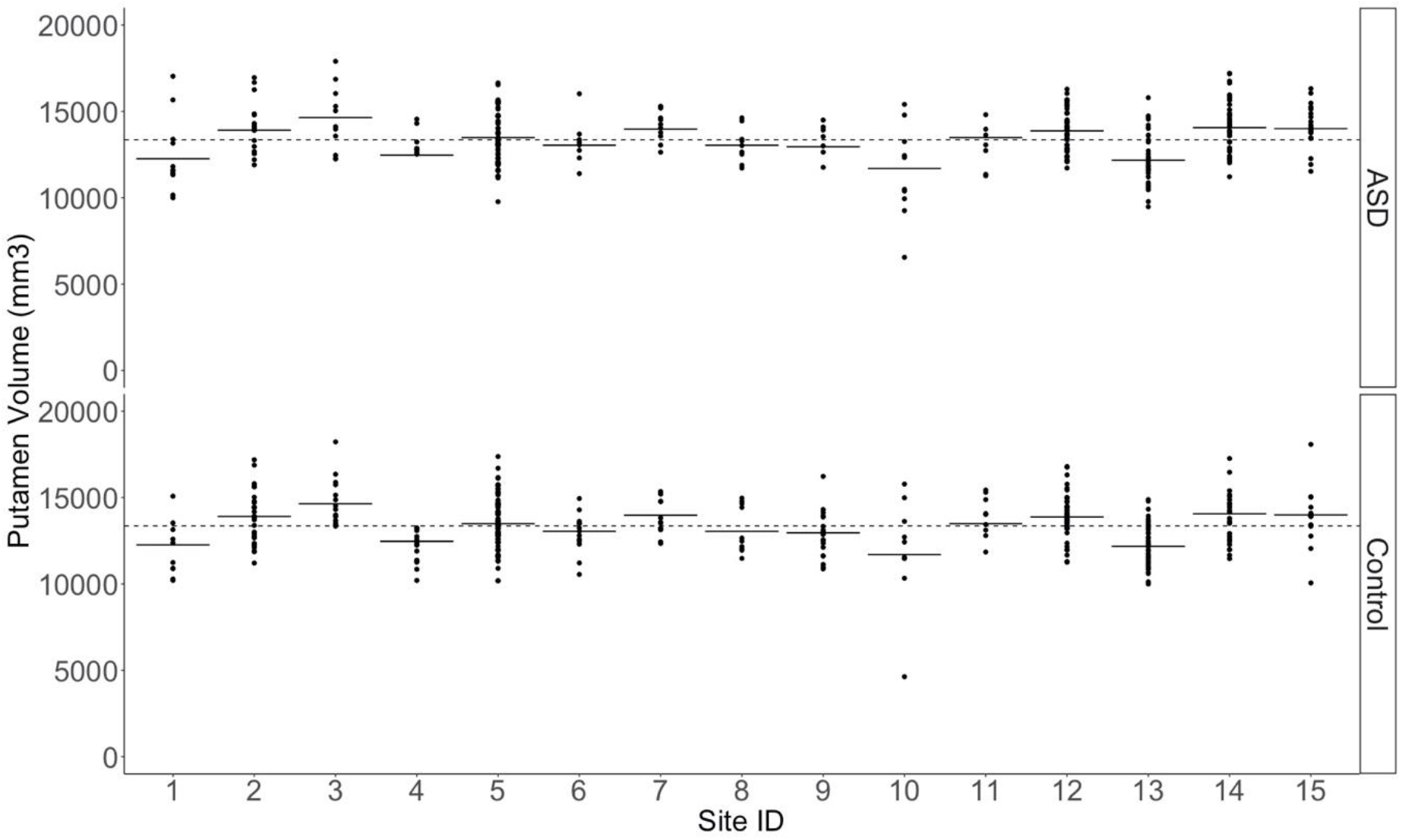
Distribution of Putamen Volumes of Autism Spectrum Disorder (ASD) and Control Participants by Scanner Site. Circles represent the brain volume of each participant. (1 = California Institute of Technology; 2 = Kenny Krieger Institute; 3 = University of Leuven Sample; 4= Ludwig Maximilians University Munich; 5 = NYU Langone Medical Center; 6 = Oregon Health and Science University; 7= Olin, Institute of Living at Hartford Hospital; 8= University of Pittsburgh School of Medicine; 9 = San Diego State University; 10 = Stanford University; 11 = Trinity Center for Health Sciences; 12 = University of California Los Angeles; 13 = University of Michigan Sample; 14 = University of Utah School of Medicine; 15 = Yale Child Study Center).

**Figures S8.**
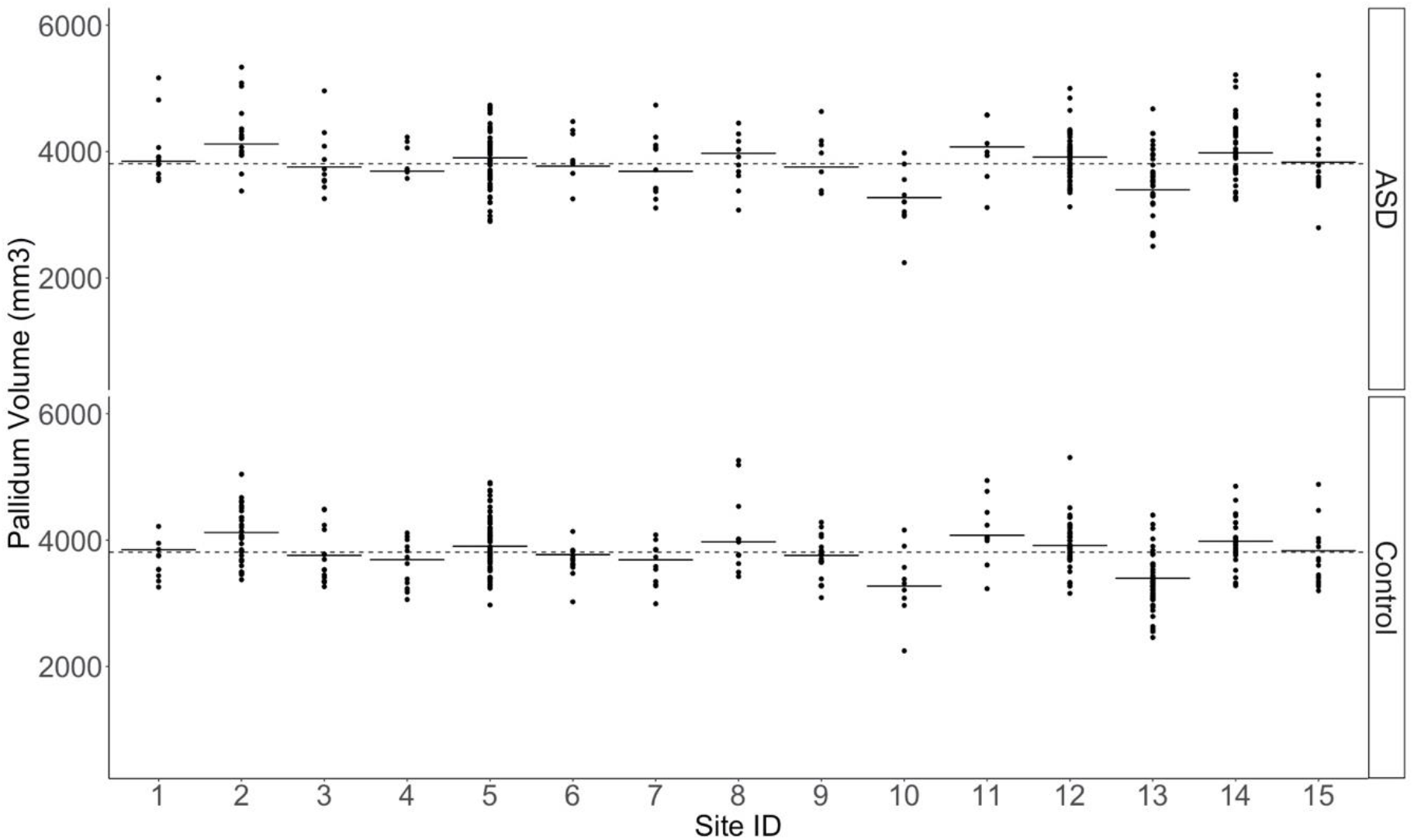
Distribution of Pallidum Volumes of Autism Spectrum Disorder (ASD) and Control Participants by Scanner Site. Circles represent the brain volume of each participant. (1 = California Institute of Technology; 2 = Kenny Krieger Institute; 3 = University of Leuven Sample; 4= Ludwig Maximilians University Munich; 5 = NYU Langone Medical Center; 6 = Oregon Health and Science University; 7= Olin, Institute of Living at Hartford Hospital; 8= University of Pittsburgh School of Medicine; 9 = San Diego State University; 10 = Stanford University; 11 = Trinity Center for Health Sciences; 12 = University of California Los Angeles; 13 = University of Michigan Sample; 14 = University of Utah School of Medicine; 15 = Yale Child Study Center).

**Figures S9.**
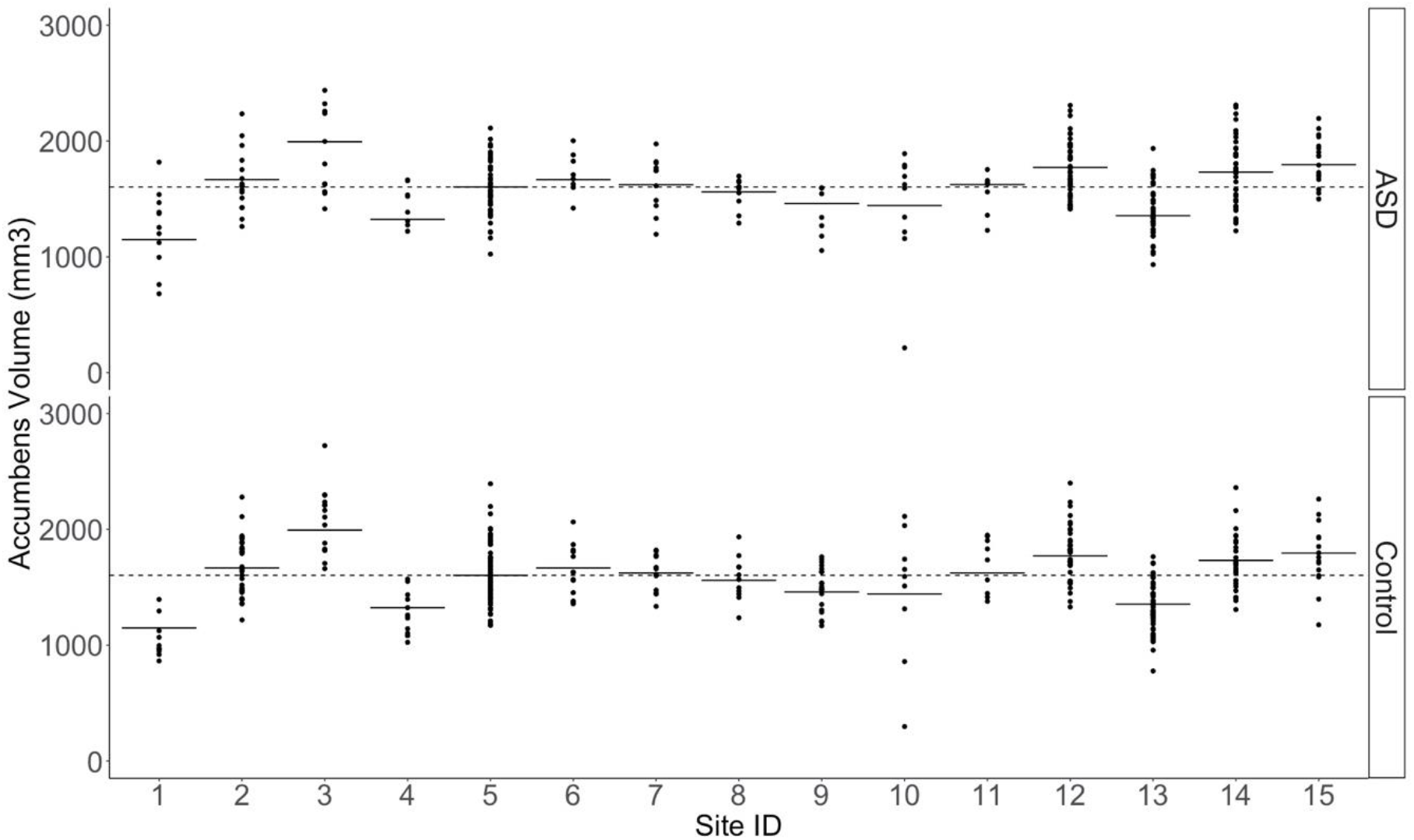
Distribution of the Accumbens Volumes of Autism Spectrum Disorder (ASD) and Control Participants by Scanner Site. Circles represent the brain volume of each participant. (1 = California Institute of Technology; 2 = Kenny Krieger Institute; 3 = University of Leuven Sample; 4= Ludwig Maximilians University Munich; 5 = NYU Langone Medical Center; 6 = Oregon Health and Science University; 7= Olin, Institute of Living at Hartford Hospital; 8= University of Pittsburgh School of Medicine; 9 = San Diego State University; 10 = Stanford University; 11 = Trinity Center for Health Sciences; 12 = University of California Los Angeles; 13 = University of Michigan Sample; 14 = University of Utah School of Medicine; 15 = Yale Child Study Center).

**Figures S10.**
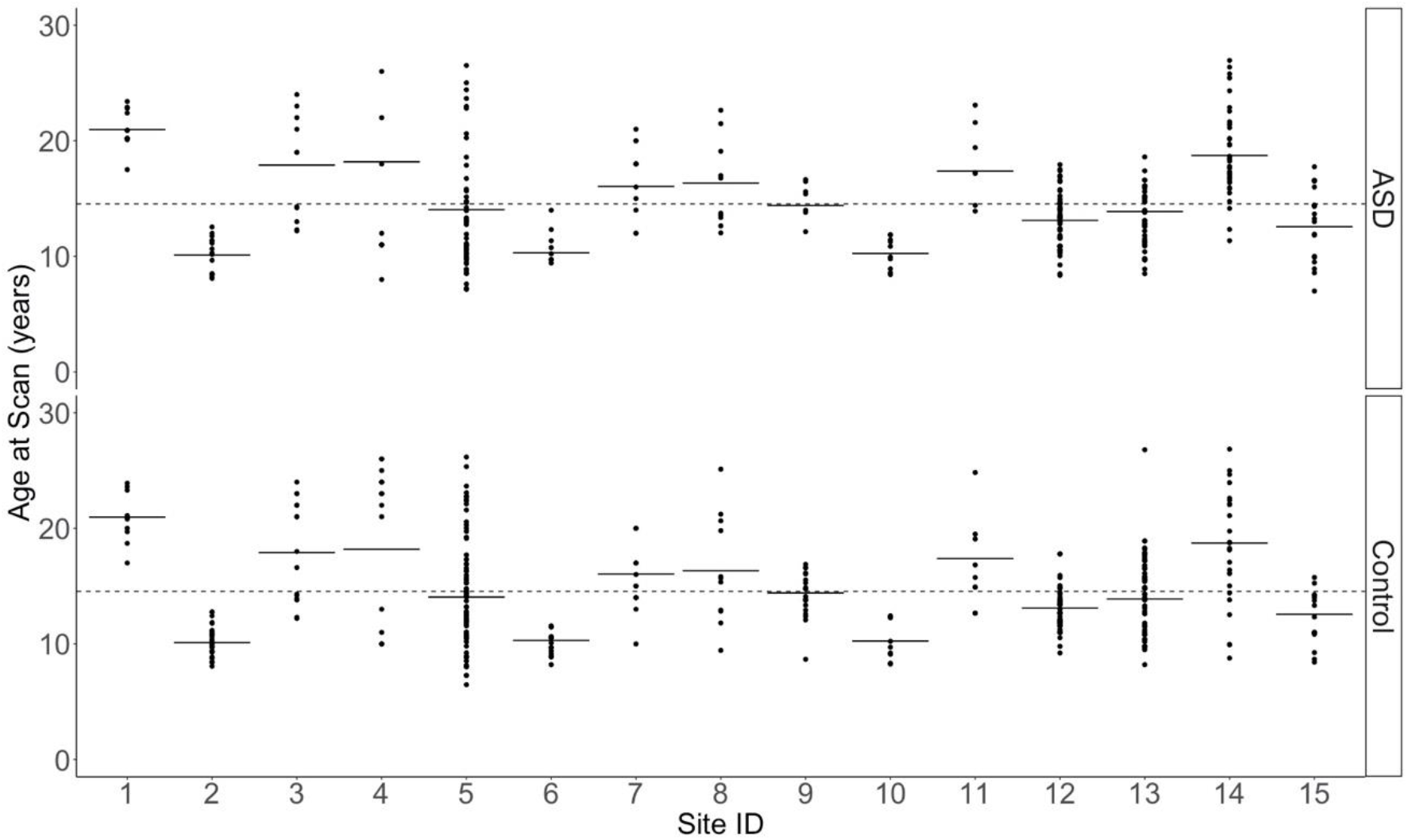
Distribution of Age of Autism Spectrum Disorder (ASD) and Control Participants by Scanner Site. Circles represent the brain volume of each participant. (1 = California Institute of Technology; 2 = Kenny Krieger Institute; 3 = University of Leuven Sample; 4= Ludwig Maximilians University Munich; 5 = NYU Langone Medical Center; 6 = Oregon Health and Science University; 7= Olin, Institute of Living at Hartford Hospital; 8= University of Pittsburgh School of Medicine; 9 = San Diego State University; 10 = Stanford University; 11 = Trinity Center for Health Sciences; 12 = University of California Los Angeles; 13 = University of Michigan Sample; 14 = University of Utah School of Medicine; 15 = Yale Child Study Center).

**Figures S11.**
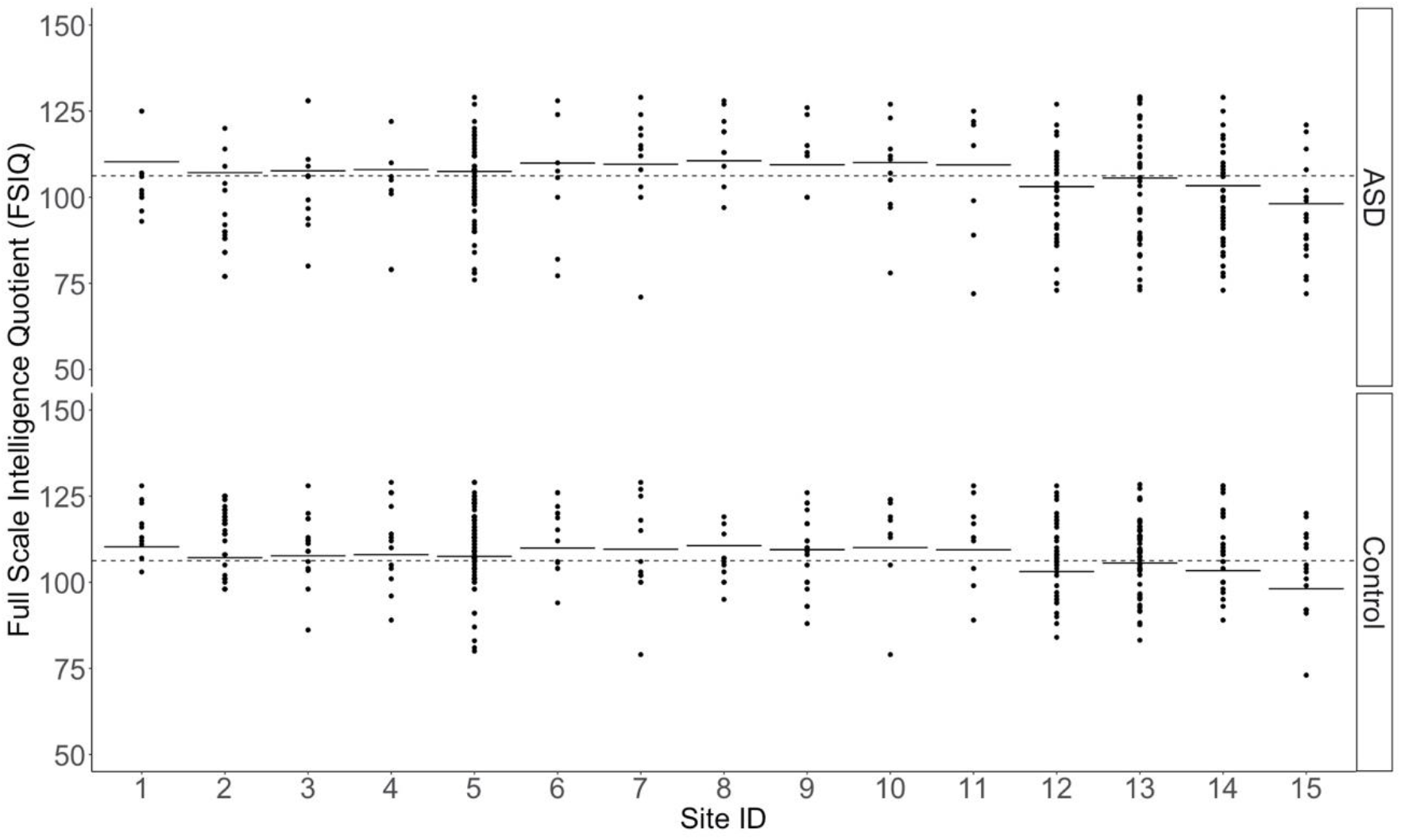
Distribution of FSIQ of Autism Spectrum Disorder (ASD) and Control Participants by Scanner Site. Circles represent the brain volume of each participant. (1 = California Institute of Technology; 2 = Kenny Krieger Institute; 3 = University of Leuven Sample; 4= Ludwig Maximilians University Munich; 5 = NYU Langone Medical Center; 6 = Oregon Health and Science University; 7= Olin, Institute of Living at Hartford Hospital; 8= University of Pittsburgh School of Medicine; 9 = San Diego State University; 10 = Stanford University; 11 = Trinity Center for Health Sciences; 12 = University of California Los Angeles; 13 = University of Michigan Sample; 14 = University of Utah School of Medicine; 15 = Yale Child Study Center).

**Figures S12.**
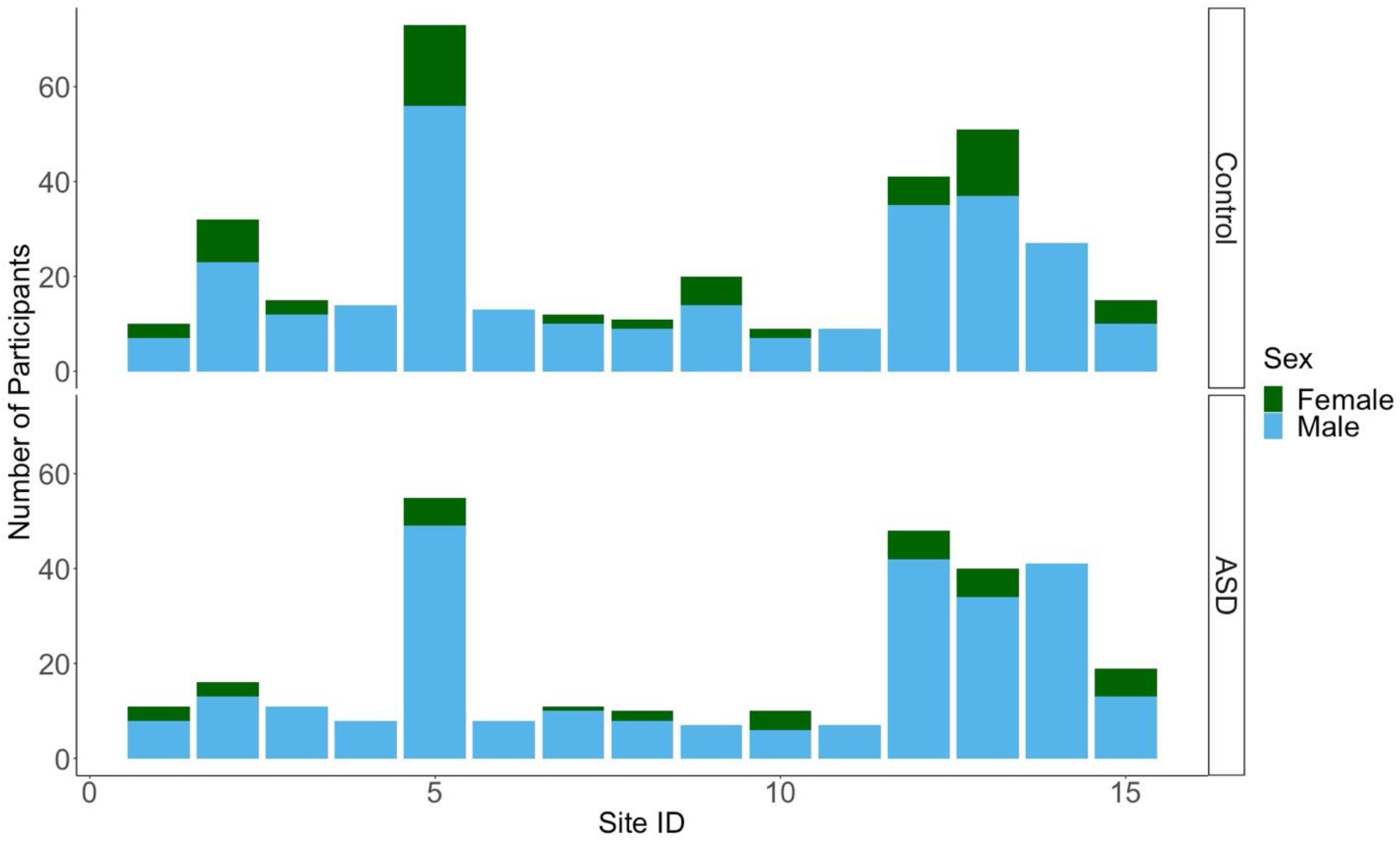
Distribution of Participants by Scanner Site, Sex, and Group (Autism Spectrum Disorder (ASD) & Controls) Circles represent the brain volume of each participant. (1 = California Institute of Technology; 2 = Kenny Krieger Institute; 3 = University of Leuven Sample; 4= Ludwig Maximilians University Munich; 5 = NYU Langone Medical Center; 6 = Oregon Health and Science University; 7= Olin, Institute of Living at Hartford Hospital; 8= University of Pittsburgh School of Medicine; 9 = San Diego State University; 10 = Stanford University; 11 = Trinity Center for Health Sciences; 12 = University of California Los Angeles; 13 = University of Michigan Sample; 14 = University of Utah School of Medicine; 15 = Yale Child Study Center).

**Figures S13.**
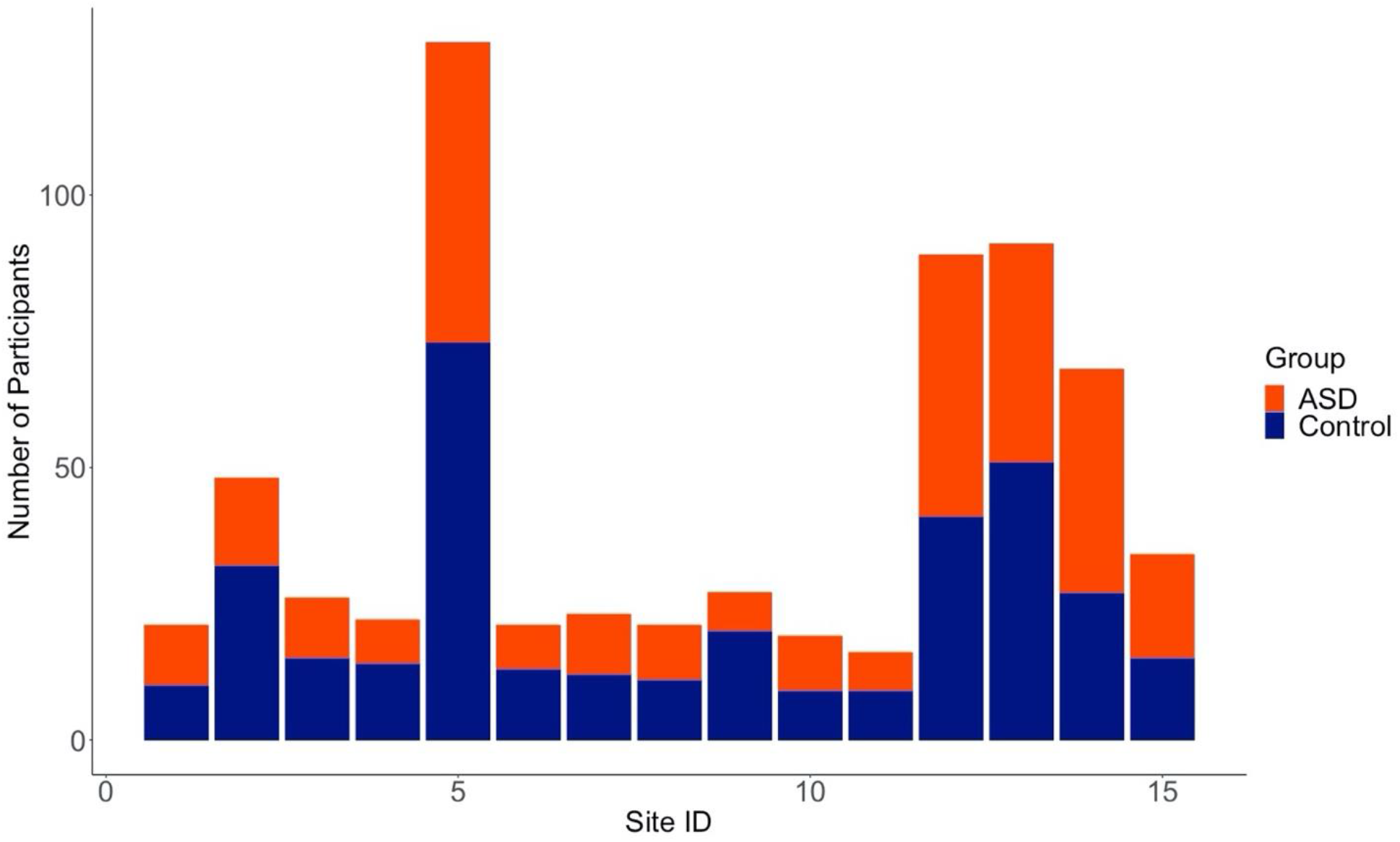
Distribution of Participants by Scanner Site and Group (Autism Spectrum Disorder (ASD) & Controls) Circles represent the brain volume of each participant. (1 = California Institute of Technology; 2 = Kenny Krieger Institute; 3 = University of Leuven Sample; 4= Ludwig Maximilians University Munich; 5 = NYU Langone Medical Center; 6 = Oregon Health and Science University; 7= Olin, Institute of Living at Hartford Hospital; 8= University of Pittsburgh School of Medicine; 9 = San Diego State University; 10 = Stanford University; 11 = Trinity Center for Health Sciences; 12 = University of California Los Angeles; 13 = University of Michigan Sample; 14 = University of Utah School of Medicine; 15 = Yale Child Study Center).

**Figures S13a.**
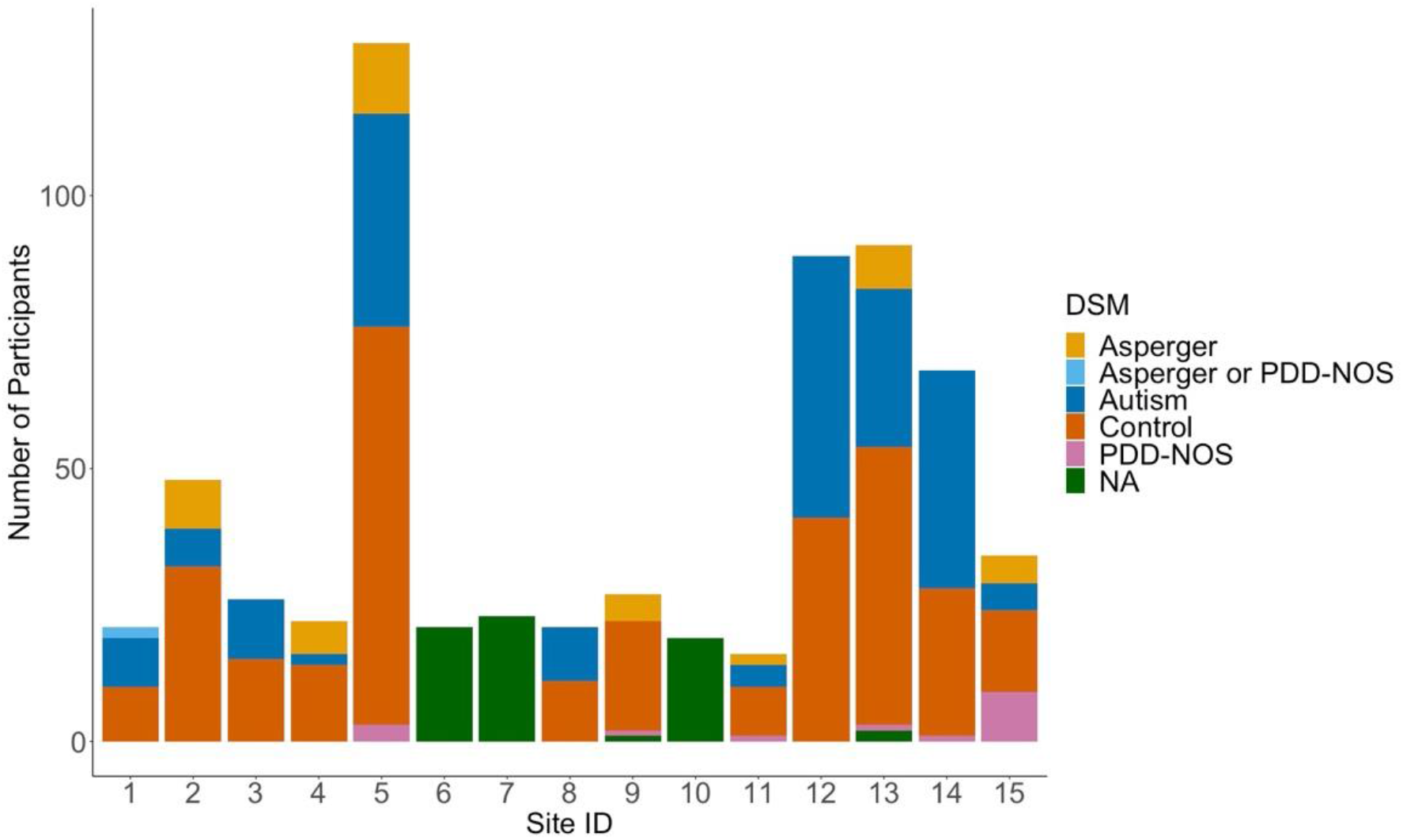
Distribution of Participants by Scanner Site and DSM-IV-TR diagnostic when available. DSM-IV-TR. fourth and text revised edition of the Diagnostic and Statistical Manual of Mental Disorder. ASD: Autism Spectrum Disorder. PDD-NOS: Pervasive Developmental Disorder-Not Otherwise Specified. NA: DSM-IV-TR not available. Circles represent the brain volume of each participant. (1 = California Institute of Technology; 2 = Kenny Krieger Institute; 3 = University of Leuven Sample; 4= Ludwig Maximilians University Munich; 5 = NYU Langone Medical Center; 6 = Oregon Health and Science University; 7= Olin, Institute of Living at Hartford Hospital; 8= University of Pittsburgh School of Medicine; 9 = San Diego State University; 10 = Stanford University; 11 = Trinity Center for Health Sciences; 12 = University of California Los Angeles; 13 = University of Michigan Sample; 14 = University of Utah School of Medicine; 15 = Yale Child Study Center).

### 2. Power Analyses

#### Multiple Group Confirmatory Factor Analysis (MGCFA)

Researchers suggest that the selection of a minimum sample size in factor analysis that provides a factor solution in agreement with the population structure from which the sample was taken depends on the number of observed variables (e.g. 22 regional volumes), factors (e.g. here, Total Brain Volume, TBV), number of variables per factor (e.g. here, 22 regional volumes), and the size of the communalities (i.e. the extent to which an observed variable correlates with the factor, which is small from 0.2 to 0.4, wide from 0.2 to 0.8, high from 0.6 to 0.8; Gaskin & Happell, 2014; Mundfrom, Shaw, & Ke, 2005).

To assess the size of the communalities, the R^2^ obtained from the lavInspect function (Rosseel, 2012) was inspected in each group and sample. While some regional volumes with very low communalities (<0.02) significantly loaded on the latent construct, the left accumbens and right amygdala did not load significantly for the ASD group in the sample of boys aged 6 to under 12 years old (R^2^ = 0.228, p = 0.185, R^2^ = 0.436, p = 0.76) and of boys with an FSIQ < median (R^2^ = 0.100, p = 0.164; R^2^ = 0.195, p = 0.055). Although the insignificance of these slopes (or loadings) suggests these regional volumes do not explain a proportion of variance in TBV in ASD individuals, these variables were maintained in the MGCFA models across samples since the right amygdala and left accumbens significantly contributed to explaining variance in TBV in controls. Considering the presence of few low communalities and that most communalities were wide but averaged around 0.4, we identified the level of communality as low to be conservative, requiring more participants. Thus, based on the simulation tables provided by Mundform and colleagues (2005), the MGCFA samples included in the current study were sufficiently large to obtain a factor solution in agreement with the population structure from which the sample was taken, as a minimum of 50 participants per group was required.

Yet, Mundform and colleagues’ (2005) criteria do not provide information on the power of our MGCFAs to detect group differences in allometric scaling (i.e. slopes or loadings) and volumes adjusted for allometric scaling (i.e. intercepts). To our knowledge, there are currently no packages developed to conduct power analyses to either detect the effect size of the group difference in slopes or intercepts with a predetermined sample or identify the number of participants required to observe a predetermined effect size of slope or intercept group differences, while including correlated residuals.

#### Linear Mixed Effects Models (LMEMs)

Power analyses for LMEMs were conducted with the simr package (Green & MacLeod, 2016). Confidence intervals were set to 95% and power estimations were based on a 1000 simulations. Power analyses were only run on significant group main effects and interactions.

### 3. MGCFA & LMEMs Assumptions

#### Univariate & Multivariate Normality

Univariate Normality was established by visually inspecting histograms of continuous variables (regional volumes, TBV, age, and FSIQ). Age and FSIQ were skewed and normal after scaling based on visual inspection of histograms. Raw volume values were normally distributed and became skewed after log10 transformation.

Univariate outliers were responsible for volumetric data non-normality based on visual comparison of brain volume histograms with and without outliers. Univariate outliers were identified separately in each group as the raw volume values that lied 1.5 times outside the interquartile range obtained from boxplots with ggplot2 (Wickham, 2016). Multivariate normality was examined in the LMEMs using Cook’s D (distance) with multivariate outliers defined as individuals with a Cook’s D 5 times greater than the mean. To correct for non-normality in the LMEMs, LMEMs were run once with outliers and once without univariate and multivariate outliers.

To correct for non-normality in MGCFAs, we ran the analyses with the MLR (Maximum Likelihood Robust) estimator provided by the lavaan package (Rosseel, 2012), which provides maximum likelihood parameters estimates with standardized errors and a chi-square test statistic that is robust to non-normality by implementing a mean adjustment.

#### Multicollinearity

Multicollinearity was examined with a Pearson’s Bivariate Correlation among all independent variables in the entire sample and in each group (Figures *S14-16*). Collinearity between observed variables is expected in the MGCFA since they are to load on the same factor. In LMEMs, multicollinearity between continuous variables was investigated with Pearson’s Bivariate Correlation and group differences in correlations investigated with the Fisher r to z transformation (Table *S5*). Although TBV and FSIQ were significantly correlated in the entire sample (r = 0.17, p = 1.14 × 10^-5^) due to its significant correlation in the control but not the Autism Spectrum Disorder (ASD) group, FSIQ and TBV were maintained together in the LMEMs since correlation was weak. Although TBV and age (linear and quadratic) were significantly correlated in the entire sample (r = 0.18, p = 4.42 × 10^-6^ and r = 0.16, p = 1.65 × 10^-5^ respectively) due to its significant correlation in the ASD but not the control group, age and TBV were maintained together in the LMEMs since correlation was weak. The same reasoning was applied for the correlation between age (linear and quadratic) and FSIQ (r = 0.10, p = 0.009 and r = 0.12, p = 0.002, respectively).

#### Heteroscedasticity

Based on visual inspection of the residual plots of our models, residuals similarly deviated from the predicted values of the LMEMs models. Although some data points appeared as outliers, homoscedasticity was generally met across LMEM models. These outliers disappeared in the LMEMs conducted without multivariate and univariate outliers.

**Figure S14.**
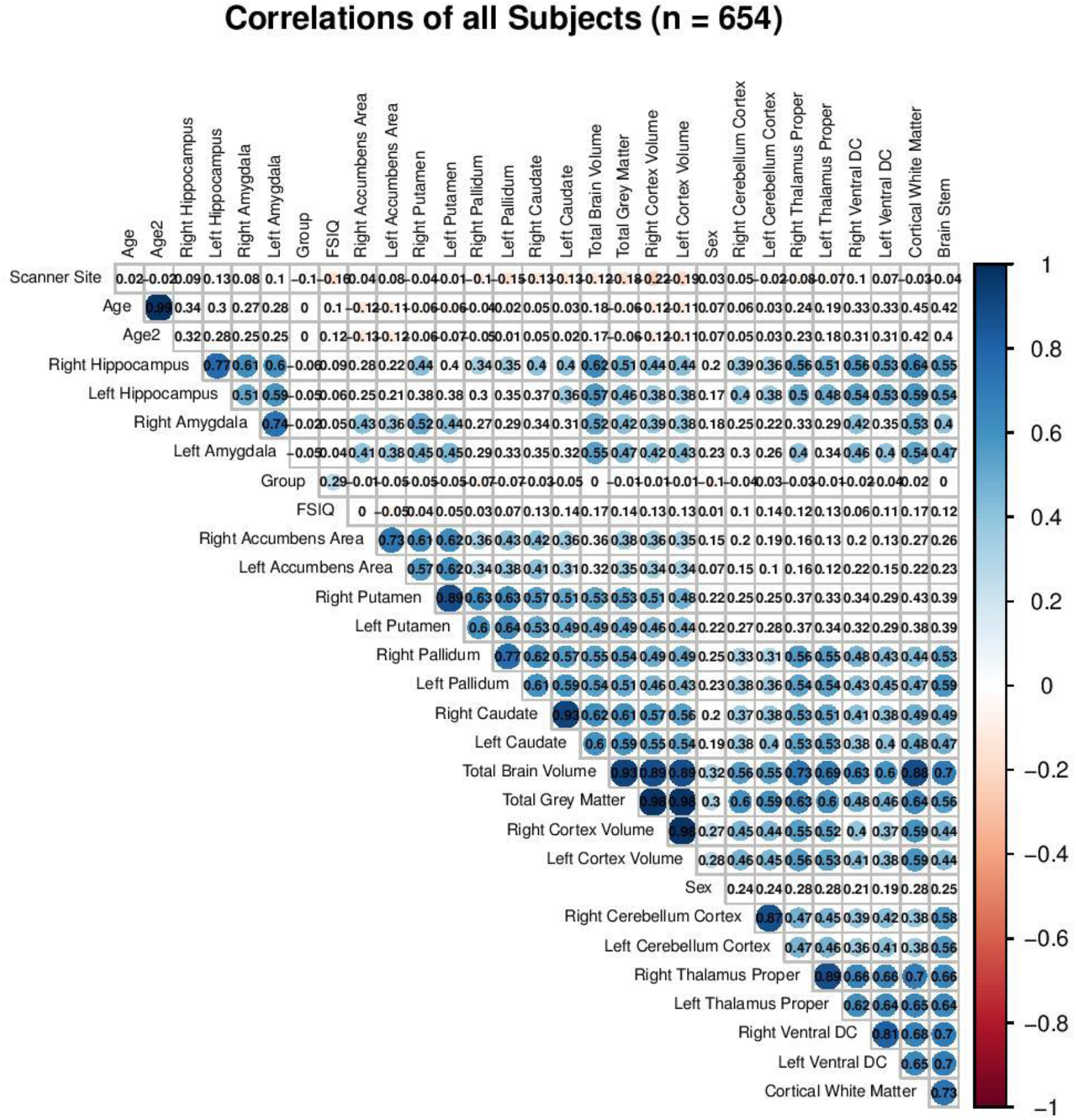
Correlation Matrix of all Cerebral Volumes, Age at Scan, Age2 (Quadratic Age), Sex, Diagnostic Group, and Full Scale Intelligence Quotient in the Entire Sample. (Correlations coefficients are in black. Significant correlations (p< 0.05) have bluer hues, indicating positive correlations, or reddish hues, indicating negative correlations. Group: Autism Spectrum Disorder or Control. DC: Diencephalon. FSIQ: Full Scale Intelligence Quotient.)”

**Figure S15.**
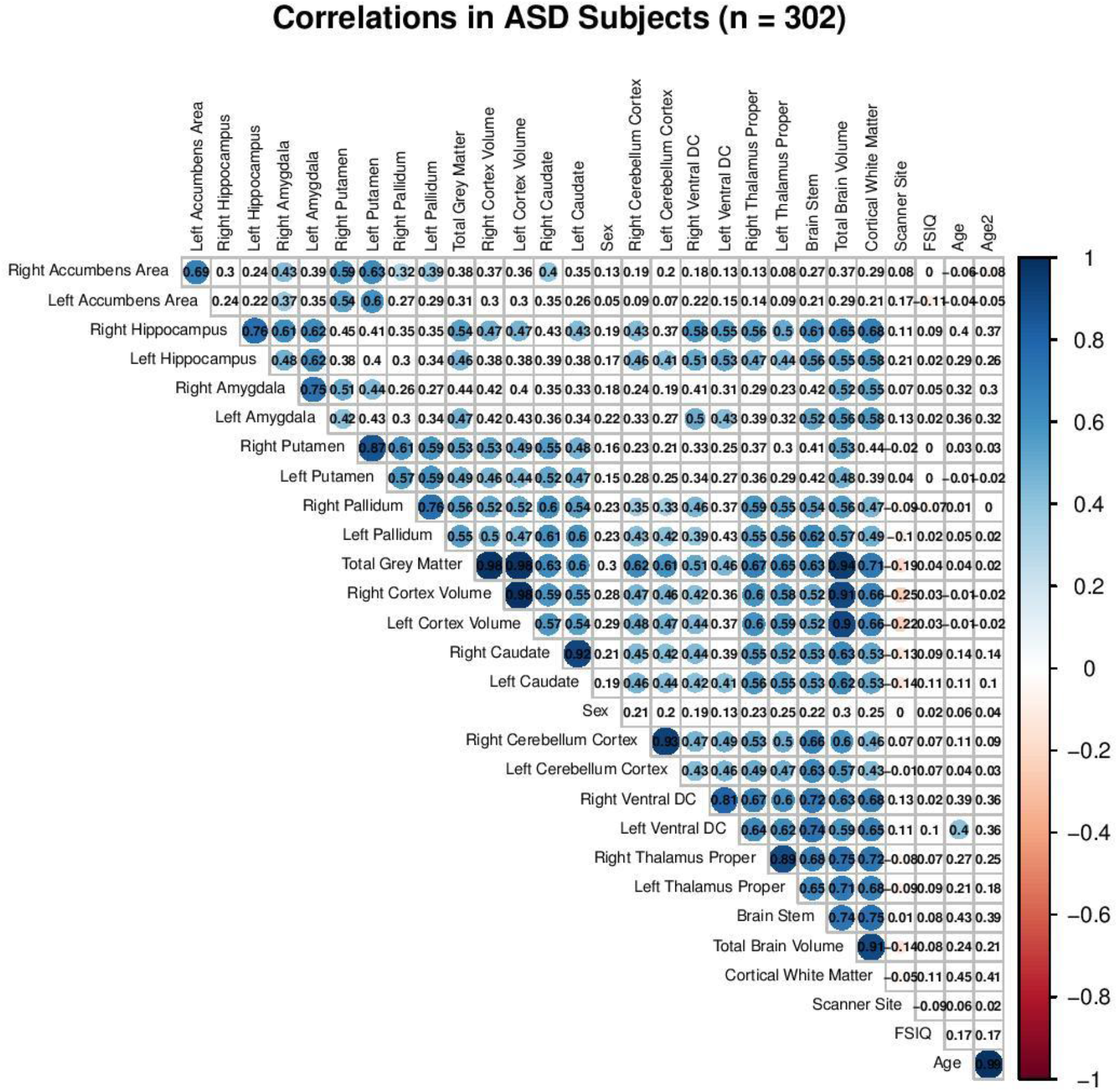
Correlation Matrix of all Cerebral Volumes, Age at Scan, Age2 (Quadratic Age), Sex, Diagnostic Group, and Full Scale Intelligence Quotient in Autism Spectrum Disorder (ASD) Participants. (Correlations coefficients are in black. Significant correlations (p< 0.05) have bluer hues, indicating positive correlations, or reddish hues, indicating negative correlations. DC: Diencephalon. FSIQ: Full Scale Intelligence Quotient.)

**Figure S16.**
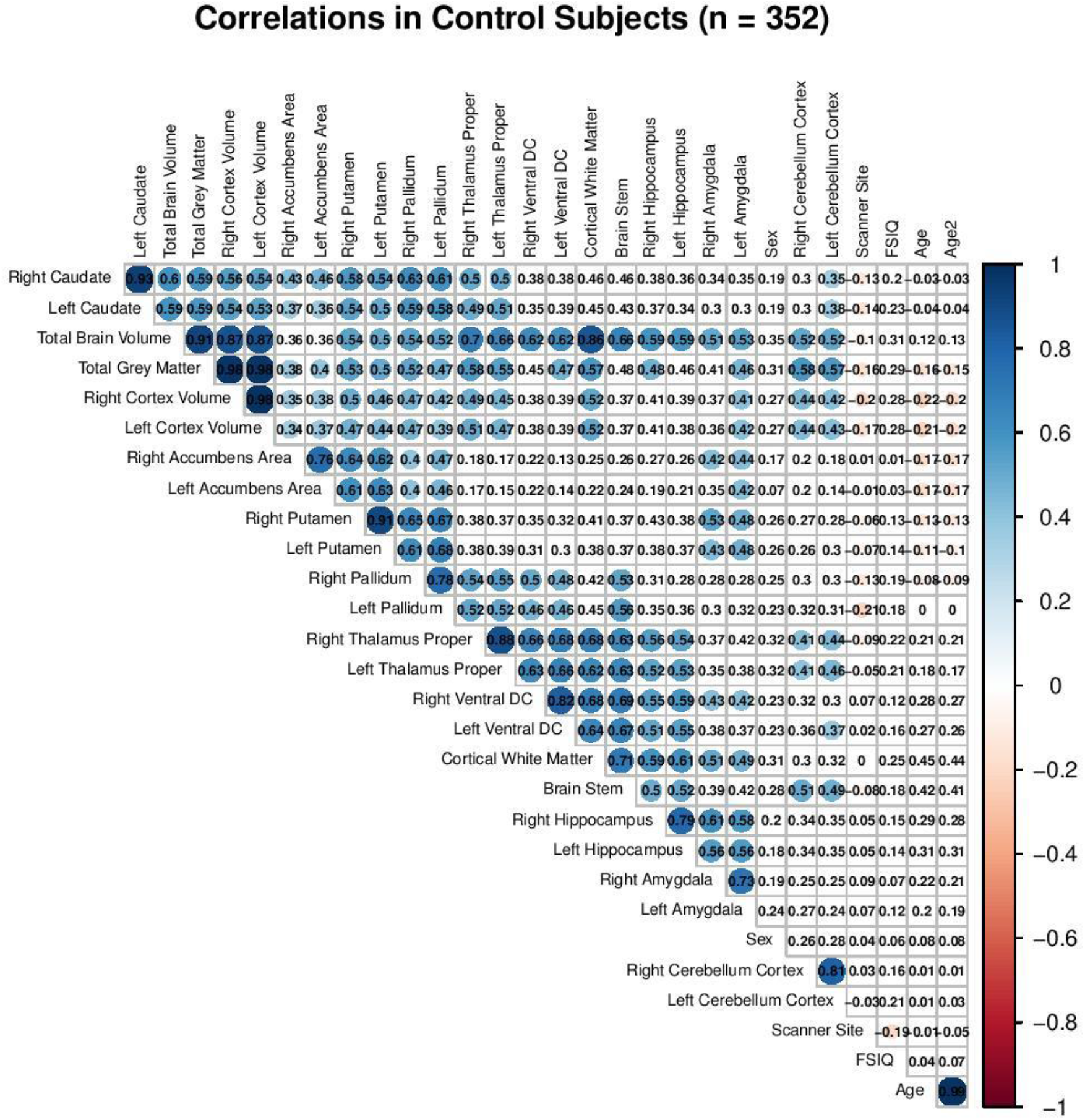
Correlation Matrix of all Cerebral Volumes, Age at Scan, Age2 (Quadratic Age), Sex, and Full Scale Intelligence Quotient in Control Participants. (Correlations coefficients are in black. Significant correlations (p< 0.05) have bluer hues, indicating positive correlations, or reddish hues, indicating negative correlations. DC: Diencephalon. FSIQ: Full Scale Intelligence Quotient.)

### 4. Replication of Zhang and Colleagues (2018): Methodology

Since the latest study examining the age and sex effects on the subcortical correlates of autism with the ABIDE I previously conducted LMEMs (1) without TBV Adjustment and (2) with linear TBV adjustment, we replicated Zhang and colleagues’ (2018) statistical analyses as well as conducted additional LMEMs (3) with allometric scaling as the TBV adjustment to validate their findings and identify if reported volumetric group differences depend on the presence and the type of adjustment for individual differences in TBV.

As in Zhang and colleagues (2018) analyses in SPSS, the dependent variables in the LMEMs were the Cortical WM Volume, Total GM Volume, the Caudate Nucleus, the Amygdala, the Hippocampus, the Thalamus, the Pallidum, the Putamen, and the Accumbens. Fixed effects differed based on the type of adjustment technique, as described below. Scanner site was always included as a random intercept and subject as a random intercept when hemisphere was included in the LMEMs model. Dependent and independent variables were entered in the models as raw values except for age (linear and quadratic), which was centered (i.e. demeaned). Significant group main effects and interactions were reported and compared across LMEMs with varying adjustment techniques. As in Zhang and colleagues (2018) p values were not adjusted for multiple comparison.

**LMEMs without TBV Adjustment**. Fixed effects were sex, age (quadratic or linear), hemisphere (except for Cortical WM and Total GM volumes), and group (ASD and Controls). Two replication strategies were put into place: a “result replication” and a “methodological replication”. In the “result replication”, models were identified based on the significant interactions reported by Zhang and colleagues (2018) to compare effect sizes even if group interactions and main effects were not significant in our sample. In the “methodological replication”, final LMEMs were identified using Zhang and colleagues’ (2018) model simplification technique, which corresponded to maintaining main effects in the model and sequentially removing all non-significant interactions (p > 0.05) from the model.

**LMEMs with linear TBV Adjustment**. As in Zhang and colleagues (2018) analyses, TBV was added as a covariate to the LMEMs identified with the “result replication” and “methodological replication” techniques. Although Zhang and colleagues (2018) ran models with and without TBV as a covariate, they commented on whether results were similar after covarying for TBV without providing any statistics (i.e. effect sizes and p values).

**Comparing LMEMs with the lack of and differing TBV Adjustment Techniques**. All brain volumes were log 10 transformed prior to scaling. LMEMs identified with the “result replication” and “methodological replication” techniques were run with the interaction of group by log10 (TBV).

### 5. MGCFA Results

#### Correlated Residuals

Correlated residuals represent correlations between observed variables not explained by the latent construct and improve model fit by allowing observe variable error terms to covary when they are part of the same factor. The correlated residuals included in the entire sample and each subsample slightly differed, since the same correlated residuals across samples did not enable acceptable model fit (Table *S14*). For instance, 76% of the correlated residuals in the boys from 6 to under 12 years old sample were included in the Entire Sample and 74% of the correlated residuals in the Entire Sample were included in the sample of boys from 6 to under 12 years old.

#### Model Fit Indices

Fit indices overall indicated good fit across samples. However, the robust TLI was slightly under < 0.95 for control boys with an FSIQ > 107.8 and the robust RMSEA was slightly over < 0.06 for ASD boys aged 6 to under 12 years old and ASD and control boys with an FSIQ > 107.8. Yet, considering that the majority of model fit indices indicated good fit, that previous studies accept fit indices values close to the model fit cutoff values (Xu & Tracey, 2017), and that adding more correlated residuals to improve fit resulted in a non-positive definite residual covariance matrix (making results not interpretable), these model fit indices were judged as acceptable (Table *S15*).

***TBV Group Differences***. In the MGCFA, TBV did not differ between individuals with and without ASD in the entire sample (ß = 0.03, SE = 0.06, p = 0.437), in boys from 6 to under 12 years old (ß = -0.08, SE = 0.10, p = 0.260), in boys from 12 to under 20 years old (ß = 0.03, SE = 0.09, p = 0.441), and in boys with an FSIQ > 107.8 (ß = -0.04, SE = 0.11, p = 0.451).

***Regional Allometric Scaling Group Differences***. Considering the exploratory result that allometric scaling differed between groups in boys from 12 to under 20 years old and in boys with an FSIQ under the median, we expected an interaction of TBV by group by sex by age and of TBV by group by sex by FSIQ in the model with TBV by group by age by sex by FSIQ as fixed effects and scanner site as random intercept in the entire sample. However, neither interaction was significant (ß = -0.03, SE = 0.20, p = 0.982, and ß = -0.07, SE = 0.25, p = 0.931, respectively).

*Boys from 12 to under 20 years old*. Table *S16.A*. shows that the robust CFI and robust RMSEA fit indices were invariant across models according to Chen’s metric invariance cutoffs (|∆CFI| >.005 & |∆RMSEA| ≥.010. Although the χ2 metric invariance test implied that groups differ in terms of allometry in the left amygdala and brain stem, the corresponding LMEMs were consistent with our interpretation of the MGCFA results suggesting the absence of an allometric scaling group difference in these volumes (respectively, ß = -0.04, SE = 0.09, p = 0.693; ß = 0.09, SE = 0.09, p = 0.322). Figure *S17* shows the allometric scaling group difference in the right hippocampus without outlier and comorbidity removal.

*Boys with an FSIQ > median (107.8)*. Table *S16.B*. shows that the robust CFI and robust RMSEA fit indices were invariant across models according to Chen’s metric invariance cutoffs (|∆CFI| >.005 & |∆RMSEA| ≥.010. The χ2 metric invariance test implied that groups also differ in terms of allometry in the left hippocampus, caudate, pallidum and ventral diencephalon. These group difference sin allometric scaling were additionally reported in the corresponding LMEMs: left hippocampus (ß = -0.29, SE = 0.10, p = 0.009), caudate (ß = -0.36, SE = 0.12, p = 0.006), pallidum (ß = -0.26, SE = 0.10, p = 0.023), and the right ventral diencephalon (ß = -0.28, SE = 0.10, p = 0.017). However, they were no longer significant after including medication as a covariate and removing outliers and comorbidities (hippocampus, ß = -0.09, SE = 0.10, p = 0.765; caudate, ß = -0.01, SE = 0.11, p = 0.429; pallidum, ß = 0.08, SE = 0.11, p = 0.607; and ventral diencephalon, ß = - 0.23, SE = 0.12, p = 0.143). Figure S18 shows the allometric scaling group difference in the left accumbens without outlier and comorbidities and Table *S18* the linear Mixed Effects Model with Quadratic Age.

**Figure S17.**
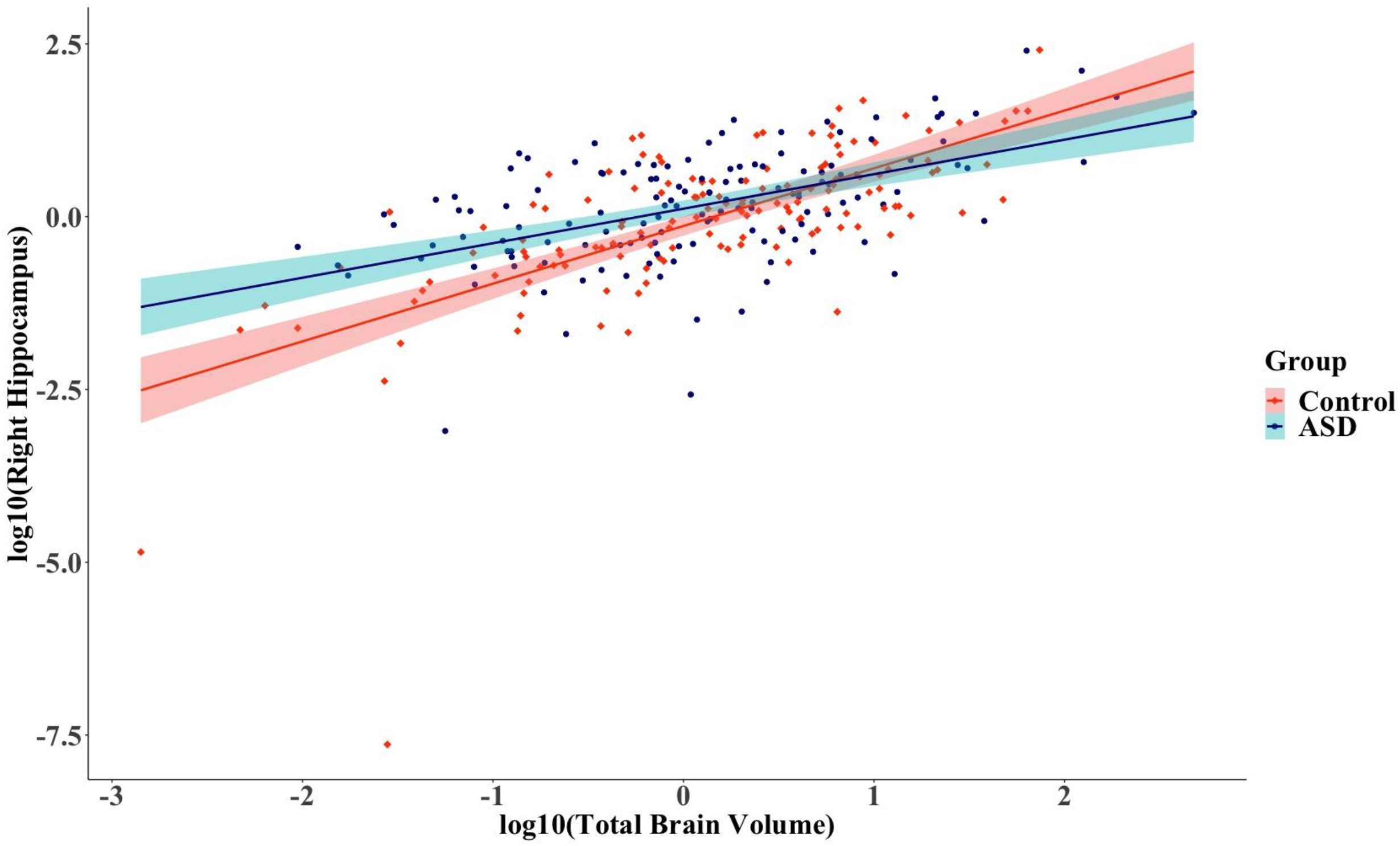
Relationship between the right hippocampus and total brain volume across groups before (N_Control_ = 141, N_ASD_= 138) outlier and comorbidity removal in boys aged 12 to under 20. ASD, Autism Spectrum Disorder. 95% confidence region are given by group. Volumes are log transformed and scaled.

**Figure S18.**
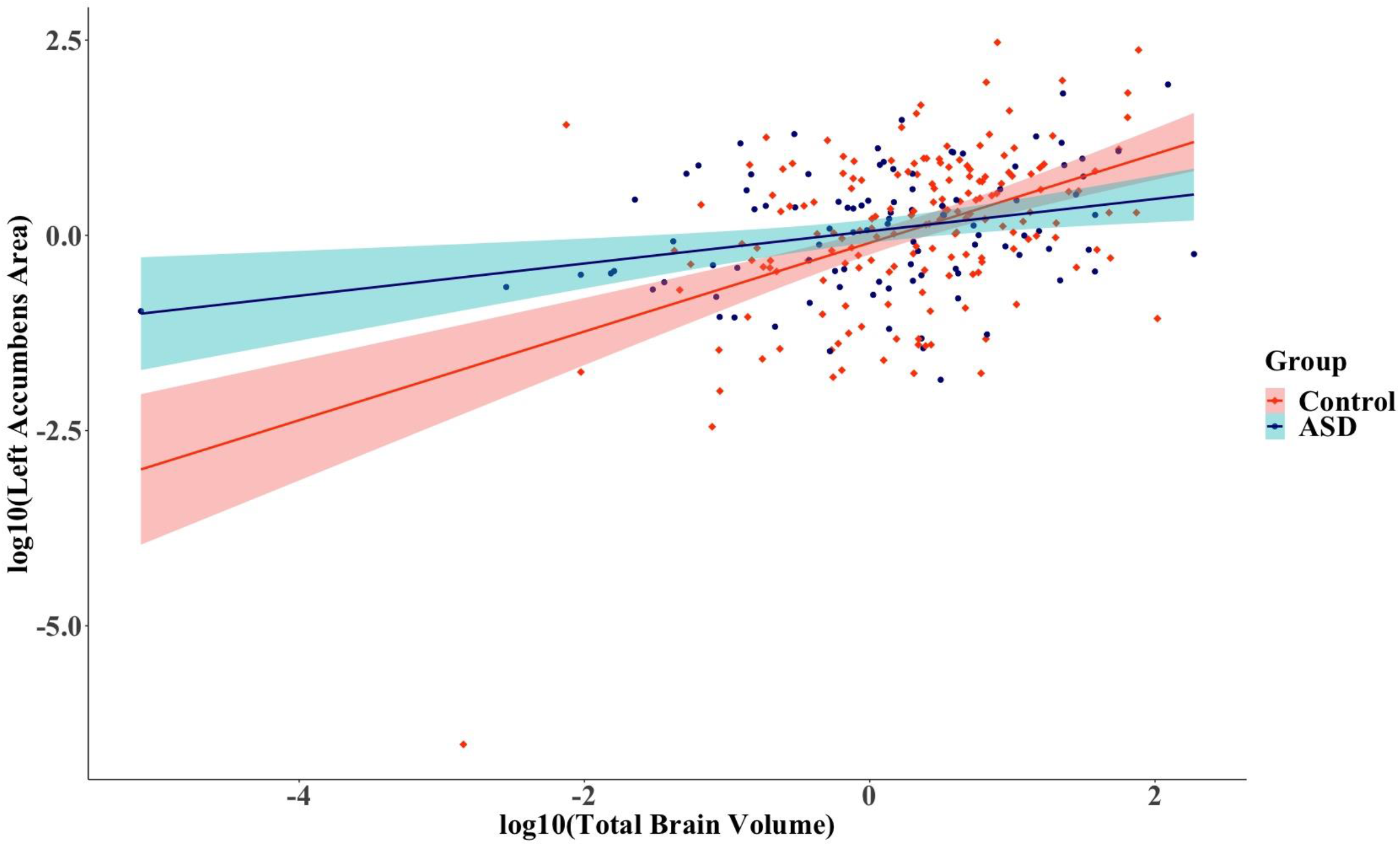
Relationship between the left accumbens and total brain volume across groups before (N_Control_ = 174, N_ASD_= 100) outlier and comorbidity removal in boys with a Full Scale Intelligence Quotient < median (107.8). ASD, Autism Spectrum Disorder. 95% confidence region are given by group. Volumes are log transformed and scaled.

### 6. Replication Results

The tables S19 to S27 compare the present study’s replication of Zhang and colleagues’ (2018) Linear Mixed Effects Models (LMEMs) without adjusting for TBV differences to Zhang and colleagues (2018) findings and to LMEMs with Linear TBV Adjustment and with Allometric Scaling TBV Adjustment. The “result replication” models were identified based on the significant interactions reported by Zhang and colleagues (2018) to compare effect sizes even if group interactions and main effects were not significant in our sample. The “methodological replication” models were identified using Zhang and colleagues’ (2018) model simplification technique, which corresponded to maintaining main effects in the model and sequentially removing all non-significant interactions (p > 0.05) from the model. Group levels were 1: Controls and 2: ASD and sex levels were 1: Female and 2: Male. Raw values were included in the models, age was centered, and scanner site and hemisphere (except for total grey volume and cortical white matter volume) were added as random intercepts.

## Notes

### Competing Interest Statement

The authors have declared no competing interest.

### Author Declarations

Data was anonymized and collected by studies approved by the regional Institutional Review Boards. Further details on participant recruitment and phenotypic and imaging data analyses are provided by Di Martino and colleagues (2014), https://doi.org/10.1038/mp.2013.78).

